# Enlarged Perivascular Spaces in the Basal Ganglia Across Epilepsy Subtypes

**DOI:** 10.1101/2024.12.31.24319825

**Authors:** Benjamin Sinclair, Clarissa Lin Yasuda, John-Paul Nicolo, Gernot Hlauschek, Thais Maria Santos Bezerra, Zhibin Chen, Lucy Vivash, Brunno Machado de Campos, Rafael Batista João, Ricardo Brioschi, Lucas Scardua-Silva, Marina Koutsodontis Machado Alvim, Patrick Kwan, Fernando Cendes, Terence J. O’Brien, Meng Law

**Author notes:** BS and CY contributed equally to this work. PK, FC, TOB and ML are joint senior authors.

## Abstract

**Introduction:** The glymphatic system is thought to be the brain’s primary waste clearance system, responsible for eliminating soluble metabolites and proteins from the central nervous system. It consists of the cerebrospinal fluid, the interstitial fluid, and a conduit between the two, perivascular spaces (PVS), which are channels formed by astroglial cells surrounding the blood vessels. PVS can be observed on high-resolution T1-weighted MRI images. Small studies have implicated PVS and glymphatics in the pathophysiology of epilepsy, potentially via reduced clearance of excitotoxic substances. This study investigates enlarged PVS burden in a large patient group with various types of epilepsy.

**Methods:** People with various types of epilepsy were recruited from the Hospital das Clínicas, Unicamp, Brazil. They were matched approximately in age and sex with healthy volunteers as controls. All participants were scanned with T1-weighted MRI on a 3T Phillips MRI scanner, resolution 1.0x1.0x1.0 mm³. A deep-learning algorithm, PINGU, was applied to segment PVS. The volumes of PVS in the White Matter (WM) and Basal Ganglia (BG) were calculated and divided by the respective volumes of WM and BG to derive the volume fractions (PVS-VF). These were used as dependent variables in a general linear model, with the diagnostic group as the independent variable of interest and age and sex included as nuisance covariates.

**Results:** We recruited 467 people with epilepsy (median age 42 years, 41.5% male), of whom 267 had temporal lobe epilepsy with hippocampal sclerosis (TLE-HS), 71 TLE with no MRI-visible lesions (TLE-NEG), 65 with focal extratemporal epilepsy (ETLE), and 64 with Idiopathic Generalized Epilepsy (IGE)). They were matched with 473 healthy controls (median age 35 years, 38.3% male). All epilepsy subtypes had higher PVS-VF in the BG compared to controls (101-140%, effect size=0.95-1.37, p<1.33x10^-15^). There was no difference in PVS-VF in the WM between the epilepsy group and healthy controls, or between different epilepsy subtypes. The TLE-HS group had an asymmetry in their PVS distribution, being larger on the contra-lateral side. This was not observed in the healthy controls or any other epilepsy subtypes. There was no association between PVS-VF and duration of illness (median duration 29 years).

**Conclusion:** Volume of PVS in the BG is enlarged in people with epilepsy. Longitudinal studies are needed to determine whether seizures have a detrimental effect on the brain’s glymphatic system, or whether impaired glymphatics contribute to the development of epilepsy.

## Introduction

Epilepsy is a highly heterogeneous group of diseases with varying etiologies, electroclinical manifestations and outcomes (Wirrell et al., 2022). The processes underlying the start and recurrence of seizures remain mostly unclear despite the numerous investigations related to diverse factors: neuroinflammation (Vezzani et al., 2019), genetic and epigenetic (Van Loo et al., 2022), among others. The extracellular environment is an important factor in the initiation, maintenance and termination of seizures (Dityatev, 2010, Leite and Peixoto-Santos, 2021). The homeostasis of the extracellular environment involves a myriad of mechanisms, among which is the glymphatic system. The glymphatic system is thought to be the brain’s primary method of removing “waste”, including degraded proteins from the interstitial space, and thus is important in the homeostasis of the extracellular environment (Mestre et al., 2020, Wardlaw et al., 2020b). Regardless of the etiology of any given epilepsy subtype, changes in the extracellular space are potentially a factor in all, as the excitability of a given neuron will be influenced by the concentration of seizure-initiating and terminating molecules in the extra-cellular space, and thus impaired glymphatics could alter a patient’s susceptibility to seizures across a range of different epilepsies. Consequently, remodelling of the extracellular matrix has been observed across a range of epilepsies, including genetic and acquired (Blondiaux et al., 2023), focal cortical dysplasia (Zamecnik et al., 2012), and mesial temporal lobe epilepsies (François et al., 2024) (Perosa et al., 2002).

One important component of the glymphatic system, the perivascular spaces (PVSs), are channels directing the flow of cerebrospinal fluid (CSF) into and out of the interstitial space. They are observable on typical clinical MRI scans (1.5T or 3T) (Wardlaw et al., 2020a). PVSs are enlarged with age (Lynch et al., 2023, Evans et al., 2023) and with various neurological diseases, including Alzheimer’s disease (Lynch et al., 2022, Ramirez et al., 2016), multiple sclerosis (Kolbe et al., 2022a), and Parkinson’s disease (Shen et al., 2021). This enlargement may be related to reduced CSF flow and impaired glymphatic functioning (Wardlaw et al., 2020a). However, the pathophysiology of PVS enlargement is not entirely understood (Christensen et al., 2021).

In this study, we aimed to characterize the PVS across a range of epilepsy subtypes in a large cohort of patients. We hypothesised that: 1) PVS would be enlarged in the white matter and basal ganglia in all epilepsy subtypes, 2) that in focal epilepsy, PVS volume would be focally altered, with a) differences with controls larger in the epileptogenic lobe of focal epilepsy and b) larger PVS around the seizure focus compared to the contralateral hemisphere, and 3) that in generalised epilepsies the PVS abnormalities would be more widespread, and not larger in any particular lobe of the brain.

## Methods

### Participants

The participants were epilepsy patients seen at the Hospital de Clínicas, Unicamp, Campinas, Brazil, between 2010 and 2022. Inclusion criteria were a diagnosis of temporal lobe epilepsy with hippocampal sclerosis (TLE-HS), temporal lobe epilepsy without visible MRI lesion (TLE-NEG), extratemporal focal epilepsy (ETLE), or idiopathic generalised epilepsy (IGE). We did not include individuals with MRIs that revealed a cerebral tumor, cerebral infarct, vascular malformations, or gliosis (from trauma or infections). Patients with multifocal epilepsy or developmental and epileptic encephalopathies were not included. In total, 267 patients with TLE-HS, 71 patients with TLE-NEG, 65 patients with ETLE, and 64 patients with IGE) were identified in the imaging database and included in this study. Additionally, 473 healthy controls (HC), consisting of members of the community in surrounding suburbs, and hospital staff were used in the study (Kerestes et al., 2024, Whelan et al., 2018).

### Image Acquisition

Participants were scanned on a 3T Philips Achieva scanner at Hospital de Clínicas, with a 3D-T1 weighted image (WI): 180 sagittal slices, TR: 7 ms, TE: 3.2 ms, slice thickness: 1.0 mm, in-plane resolution: 1.0 mm x 1.0 mm and FOV: 240x240 mm² (Whelan et al., 2018).

### Image Processing

Perivascular spaces were segmented using the Perivascular-space Identification Nnunet for Generalised Usage (PINGU) algorithm (Sinclair et al., 2024) (Figure 1). To localise the PVS to anatomical regions, Freesurfer 7.1.1 (Fischl, 2012) segmentation was run, and the WM and BG extracted (Figure 2). The WM was further subdivided into Frontal, Parietal, Temporal and Occipital WM. The BG consisted of the regions: thalamus, caudate, putamen, pallidum, accumbens and ventral diencephalon. Since the Freesurfer segmentation of WM includes the WM tracts passing through the BG and does not distinguish cortical vs subcortical WM, a separate atlas (JHU DTI 81 tract atlas (Mori, 2005)) was used to define the WM tracts passing through the BG: anterior and posterior limbs of the internal capsule, and the external capsule, and assign them to the BG region (and remove them from the whole WM region). To do this, the MNI template provided by FSL (Jenkinson et al., 2012) was non-linearly registered to each T1w image using ANTS Syn registration (Avants et al., 2008), and the deformation parameters applied to the JHU anatomical atlases.

**Figure 1:**
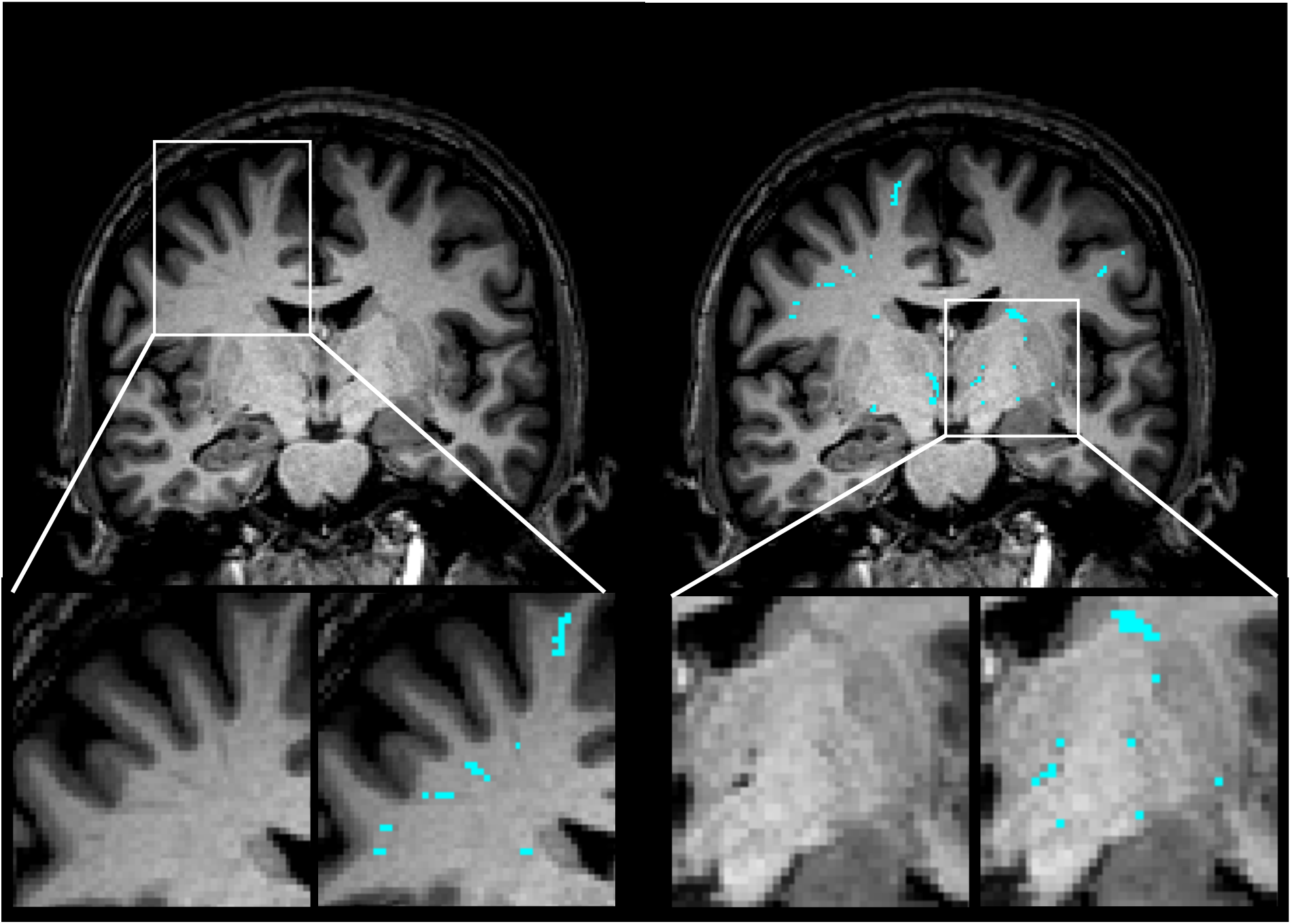
Raw T1w MRI image (left) and overlay of PINGU PVS segmentation (right) for a single patient with TLE.

**Figure 2:**
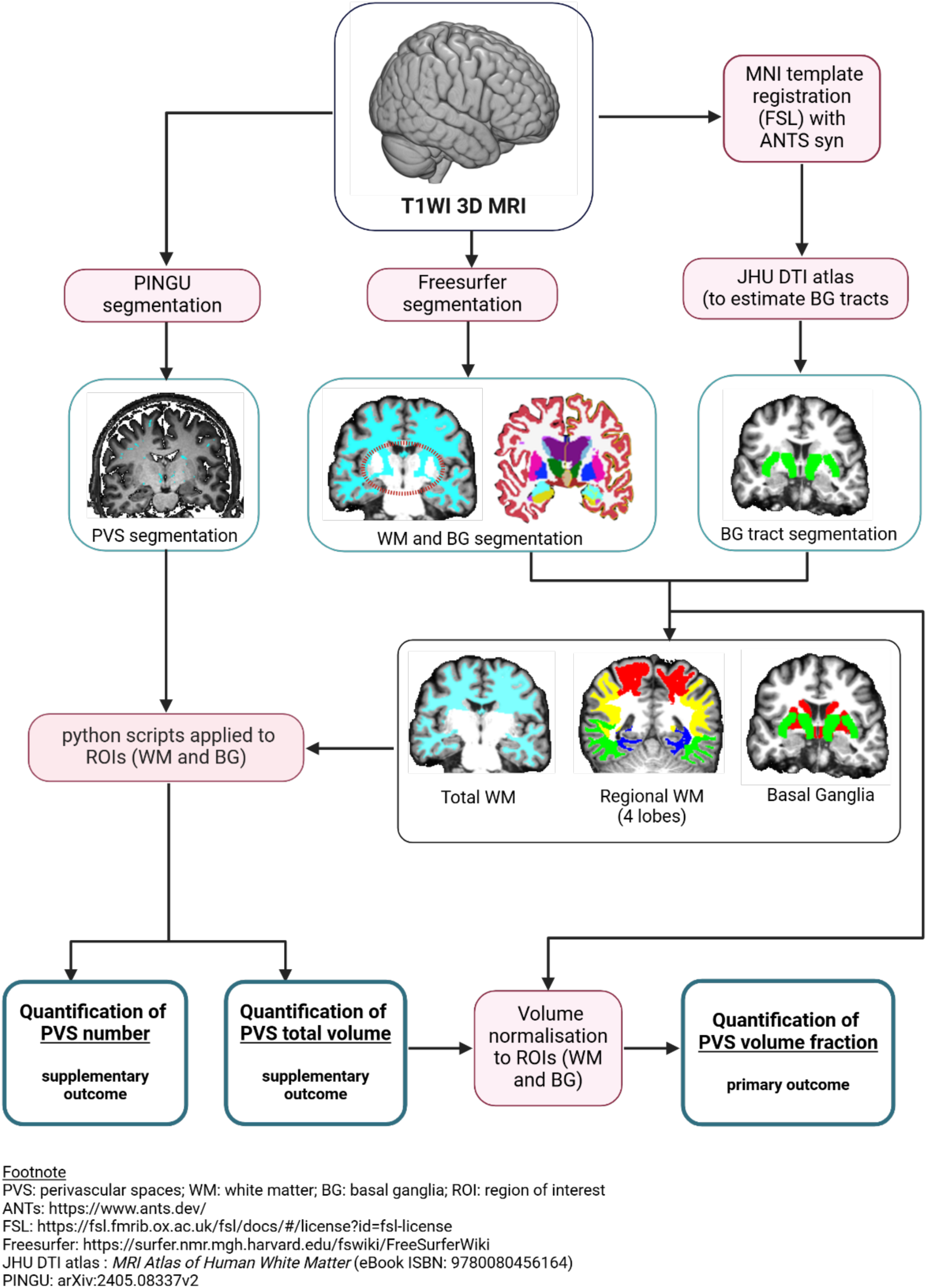
Workflow of MRI processing steps (Created in: https://BioRender.com)

Quantification of the PVS volumes and numbers in each region of interest was conducted using a Python script that masked the PVS segmentation image by each region of interest in the registered atlases. The volume of PVS was then normalised to the volume of that region for each patient to give volume fractions (VF). The primary outcome variables for each hypothesis tested were (a) PVS-VFs and two supplementary outcome measures: (b) PVS-Volume: total volumes without correction for region volume, to see whether results were robust to volume normalisation, and (c) PVS-Number, calculated as the number of distinct un-connected clusters of neighbouring PVS voxels, calculated using skimage’s measure.label algorithm, to compare with prior work and manual counting methods (Potter et al., 2015a, Potter et al., 2015b).

### Global PVS Burden

Outcome measures in the whole WM and whole BG were compared between each epilepsy subtype and healthy controls, and each epilepsy subtype to each-other. We hypothesised that each epilepsy group would have enlarged PVS compared to controls in both whole WM and whole BG. The between-epilepsy group analysis was exploratory, and no prior hypothesis was made regarding the direction of any changes.

### Sub-Regional PVS Burden

Outcome measures in the WM of each lobe (frontal, parietal, temporal, occipital) and in the thalami and “BG excluding thalami” were compared between each epilepsy type and healthy controls, and each epilepsy type to each-other. We hypothesised that TLE-HS and TLE-NEG would have enlarged PVS in the temporal lobe, ETLE would have enlarged PVS outside the temporal lobes, and IGE would have PVS enlargement across the brain. Given the importance of thalamocortical circuits in seizure propagation across a range of epilepsies and the finding of enlarged PVS in the thalamus in a small focal epilepsy group [CITE Langan], we hypothesised that the effect size of the PVS enlargement in the thalami would be greater than that of the BG excluding thalami.

### Asymmetry

For each outcome measure, the asymmetry between ipsilateral and contra-lateral hemispheres was calculated as:

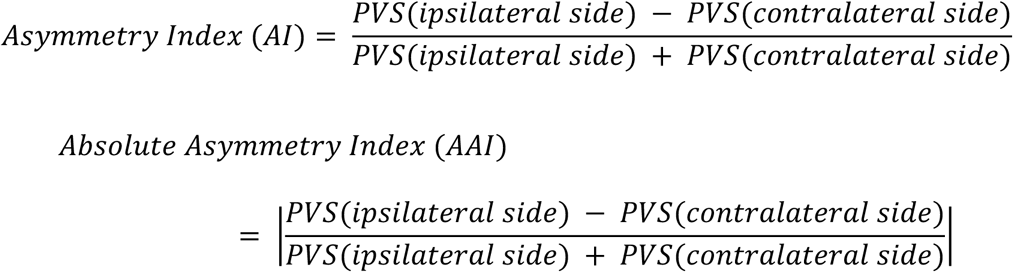

The asymmetry index ranges between −1 to 1, with 1 indicating that all PVS are on the ipsilateral side, and −1 indicating that they are all on the contra-lateral side.

We hypothesised that the focal epilepsies would have asymmetries significantly greater than zero in the whole WM and greater than in the control group. We also hypothesised that the absolute asymmetries would be more significant in the focal epilepsy groups than in the control and IGE groups. We hypothesised that asymmetry would not significantly differ from zero in the IGE group.

We further hypothesised that asymmetries would be localised to the lobe of epilepsy, i.e. that TLE-HS and TLE-NEG would have significant asymmetry in the temporal lobe WM, ETLE would have substantial asymmetry in the non-temporal lobe WM, and that all of these asymmetries in the focal epilepsies would be significantly different to controls.

### Statistics

Univariate pairwise comparisons between groups were conducted using the two-tailed Dunn’s test for each outcome measure. For each outcome measure, a general linear model (GLM) was constructed with the PVS measure as the dependent variable, group as the independent variable of interest, and age and sex included as nuisance covariates. Since some PVS measures were normally distributed, and others were skewed with long tails, two families of GLM were fit to each measure: a Gaussian family with an identity link function and a Gamma family with an inverse link function. The best-fitting model was assessed via Akaike’s information criterion (AIC), and the parameters of the best model were presented. After correction for nuisance covariates, standardised effect sizes for the Gaussian models were calculated using R’s emmeans package (the effect size of a gamma model with an inverse link function is not readily interpretable).

Association of PVS measures with duration of illness was tested with a separate GLM, with duration of illness as the independent variable of interest, and age and sex as nuisance covariates. All epilepsy patients (n=467) were included in this test, and the type of epilepsy was not considered in the model.

## Results

### Participants

Participant demographics are reported in Table 1. The TLE-HS and TLE-NEG groups were older than the control, ETLE and IGE groups, and the ETLE and IGE groups were younger than the control group (Table S1).

**Table 1:** Cohort Demographics.

| Group | n | Age | Duration (years) | Sex |
| --- | --- | --- | --- | --- |
|  |  | median (IQR) | median (IQR) | (M%) |
| Control | 473 | 35.5 (24) | NA | 38.3 |
| TLE-HS | 268 | 46 (12) | 35 (19) | 38.6 |
| TLE-NEG | 71 | 46 (18.5) | 23.5 (21) | 39.4 |
| ETLE | 65 | 31 (16) | 20 (17) | 60.0 |
| IGE | 64 | 33 (13) | 21 (15) | 37.5 |

### Global PVS Burden

In the WM, no epilepsy group had significantly different PVS-VF (Table 2, Figure 3, Table S2.1) or PVS-Number (Table S2.3) than controls. The IGE group had significantly lower PVS-VF than the TLE-HS group (−32.1%, p = 0.009) and the TLE-NEG group (−35.6%, p = 0.039), but these differences were not significant after correction for age and sex in the GLMs (Table 2), indicating that the between group differences were driven by the older age of the TLE-HS and TLE-NEG groups.

**Figure 3:**
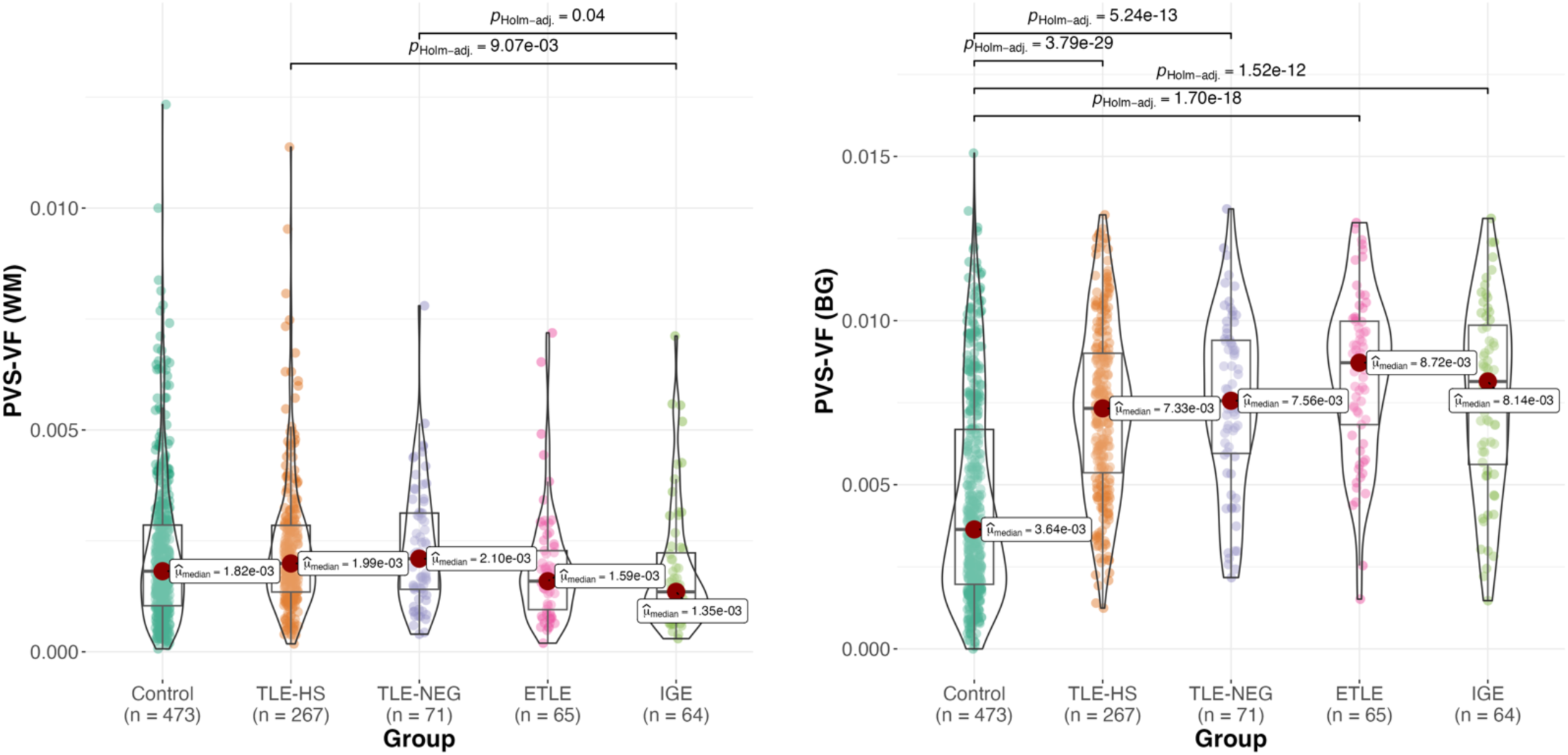
PVS volume fraction in white matter (WM, left) and Basal Ganglia (BG, right) by group. Footnote: TLE: temporal lobe epilepsy; HS: hippocampal sclerosis; TLE-NEG: temporal lobe epilepsy with MRI negative; ETLE: extratemporal epilepsy; IGE: Idiopathic generalised epilepsy

**Table 2:** Group differences in PVS volume fraction in the total white matter.

| Group |  | median (IQR) |  | Univariate |  |  | General Linear Model |  |  |  |
| --- | --- | --- | --- | --- | --- | --- | --- | --- | --- | --- |
| G1 | G2 | G1 ($\times 10^{-2}$) | G2 ($\times 10^{-2}$) | $\Delta\%$ | Dunn | p-Holm | beta (sderr) | d | z | p-Tukey |
| Control | TLE-HS | 0.18(0.18) | 0.20(0.15) | 9.6 | 2.26 | 0.168 | -4.45(22.80)$\times 10+0$ | -0.032 | -0.20 | 1.000 |
| Control | TLE-NEG | 0.18(0.18) | 0.21(0.17) | 15.5 | 1.60 | 0.442 | 4.11(37.20)$\times 10+0$ | -0.039 | 0.11 | 1.000 |
| Control | ETLE | 0.18(0.18) | 0.16(0.13) | -12.4 | 1.31 | 0.570 | 1.43(4.85)$\times 10+1$ | -0.033 | 0.30 | 0.998 |
| Control | IGE | 0.18(0.18) | 0.14(0.13) | -25.6 | 2.17 | 0.170 | 3.86(4.95)$\times 10+1$ | -0.109 | 0.78 | 0.93 |
| TLE-HS | TLE-NEG | 0.20(0.15) | 0.21(0.17) | 5.4 | 0.23 | 1.000 | 8.56(38.80)$\times 10+0$ | -0.006 | 0.22 | 0.999 |
| TLE-HS | ETLE | 0.20(0.15) | 0.16(0.13) | -20 | 2.50 | 0.099 | 1.88(4.99)$\times 10+1$ | 0.000 | 0.38 | 0.995 |
| TLE-HS | IGE | 0.20(0.15) | 0.14(0.13) | -32.1 | 3.32 | 0.009 | 4.30(5.08)$\times 10+1$ | -0.077 | 0.85 | 0.907 |
| TLE-NEG | ETLE | 0.21(0.17) | 0.16(0.13) | -24.1 | 2.19 | 0.170 | 1.02(5.82)$\times 10+1$ | 0.006 | 0.18 | 1.000 |
| TLE-NEG | IGE | 0.21(0.17) | 0.14(0.13) | -35.6 | 2.86 | 0.039 | 3.44(5.90)$\times 10+1$ | -0.071 | 0.58 | 0.975 |
| ETLE | IGE | 0.16(0.13) | 0.14(0.13) | -15.1 | 0.66 | 1.000 | 2.42(6.53)$\times 10+1$ | -0.077 | 0.37 | 0.995 |

TLE-HS: temporal lobe epilepsy with hippocampal sclerosis; IGE: idiopathic generalised epilepsy;
TLE-NEG: temporal lobe epilepsy with MRI negative; ETLE: extratemporal epilepsy

In the basal ganglia, all epilepsy groups had larger PVS-VF (Table 3, Figure 3, Table S3.1) and PVS-Number (Table S3.3) than the control group, TLE-HS (101.4%, p < 1x10^-20^), TLE-NEG (107.8%, p=5.24x10^-13^), ETLE (139.6%, p = 1.70x10^-18^), IGE (123.9%, p = 1.52x10^-12^). These findings were all significant after correction for age and sex and had large standardised effect sizes: TLE-HS: 0.951, TLE-NEG: 1.039, ETLE: 1.367, IGE: 1.108. Additionally, after correcting for age and sex, the ETLE group had larger PVS-VF than the TLE-HS group (d = 0.416, p = 0.028).

**Table 3:** Group differences in PVS volume fraction in the Basal Ganglia.

| Group |  | median (IQR) |  |  | Univariate |  | General Linear Model |  |  |  |
| --- | --- | --- | --- | --- | --- | --- | --- | --- | --- | --- |
| G1 | G2 | G1 (x10<sup>-2</sup>) | G2 (x10<sup>-2</sup>) | Δ% | Dunn | p-Holm | beta (sderr) | d | z | p-Tukey |
| Control | TLE-HS | 0.36(0.47) | 0.73(0.36) | 101.4 | 11.41 | <1x10-20 | 27.40(2.24)x10-4 | 0.951 | 12.22 | <1x10-20 |
| Control | TLE-NEG | 0.36(0.47) | 0.76(0.34) | 107.8 | 7.5 | 5.24x10-13 | 30.00(3.69)x10-4 | 1.039 | 8.11 | 3.77x10-15 |
| Control | ETLE | 0.36(0.47) | 0.87(0.32) | 139.6 | 9.02 | 1.70x10-18 | 39.40(3.88)x10-4 | 1.367 | 10.15 | <1x10-20 |
| Control | IGE | 0.36(0.47) | 0.81(0.42) | 123.9 | 7.34 | 1.52x10-12 | 31.90(3.86)x10-4 | 1.108 | 8.27 | 1.78x10-15 |
| TLE-HS | TLE-NEG | 0.73(0.36) | 0.76(0.34) | 3.2 | 0.61 | 1.000 | 2.56(3.85)x10-4 | 0.089 | 0.66 | 0.961 |
| TLE-HS | ETLE | 0.73(0.36) | 0.87(0.32) | 19.0 | 2.31 | 0.124 | 12.00(4.13)x10-4 | 0.416 | 2.90 | 0.028 |
| TLE-HS | IGE | 0.73(0.36) | 0.81(0.42) | 11.2 | 0.75 | 1.000 | 4.53(4.10)x10-4 | 0.157 | 1.11 | 0.79 |
| TLE-NEG | ETLE | 0.76(0.34) | 0.87(0.32) | 15.3 | 1.39 | 0.819 | 9.44(5.07)x10-4 | 0.327 | 1.86 | 0.32 |
| TLE-NEG | IGE | 0.76(0.34) | 0.81(0.42) | 7.8 | 0.13 | 1.000 | 1.97(5.04)x10-4 | 0.068 | 0.39 | 0.995 |
| ETLE | IGE | 0.87(0.32) | 0.81(0.42) | -6.6 | 1.23 | 0.881 | -7.46(5.10)x10-4 | -0.259 | -1.46 | 0.565 |

TLE-HS: temporal lobe epilepsy with hippocampal sclerosis; IGE: idiopathic generalised epilepsy;
TLE-NEG: temporal lobe epilepsy with MRI negative; ETLE: extratemporal epilepsy

Without volume correction (Tables S2.2, S3.2), the only between-group difference in the WM was a greater volume of PVS in TLE-HS compared to IGE, again, not significant after correction for age and sex, whilst all between-group differences in the BG remained significant without volume correction (Table S3.2

### Sub-Regional PVS Burden

In the frontal lobe WM, TLE-HS and TLE-NEG had significantly larger PVS-VF (Table S2.1), and number (Table S2.3), than Controls, IGE and ETLE, though none of these associations were significant after correction for age and sex. In the temporal lobe, the IGE group had smaller PVS-VF than Controls, TLE-HS and TLE-NEG, though not significant after correction for age and sex. In the occipital lobe, all groups had significantly lower PVS-VF and number than the control group, significant with age and sex correction and without correction for volume. However, the distributions of PVS in this lobe were overwhelmingly driven by the number of patients with zero PVS (Table S2.3, Figure S1).

In the BG excluding thalami, all epilepsy groups had significantly larger PVS-VF than controls. However, in the thalami, only the TLE-HS group had significantly larger PVS-VF than controls, which was not significant after age and sex correction (Table S3.1). Again, the distribution of PVS in the thalamus was heavily skewed towards zero (Table S3.3, Figure S2).

### PVS Asymmetry

The only group with a significantly non-zero PVS-VF asymmetry in the WM was the TLE-HS group (asym = −2.4(1.7)x10^-2^, p = 0.006, Table S4.0.1), which had a significantly negative asymmetry in all lobes other than the occipital lobe (indicating more PVS on the contra-lateral side). The lobe with the largest asymmetry in the TLE-HS group was the temporal lobe (−9.3 (7.1) x10^-2^, p=0.011). However, the control group also had a significantly negative asymmetry (left hemisphere - right hemisphere) in the temporal lobe (−6.8(5.3) x10^-2^, p = 0.011), though not in the whole WM, and the distribution of asymmetries in the temporal lobe was highly skewed, with asymmetries of −1, 0 and 1 dominating the distribution (Figure S3). Further, the TLE-HS group also had a significantly negative asymmetry, albeit smaller, in the frontal (−3.7 x10^-2^ (p = 0.023)) and parietal (−3.1x10^-2^ (p = 0.035)) lobes (which have more normally distributed asymmetry values, Figure S3). No other epilepsy group had significantly non-zero asymmetry in any lobe other than IGE in the Frontal lobe.

There were no differences between groups in PVS-VF asymmetry in the WM (Table S4.1, Figure 4). Neither the TLE-HS nor the TLE-NEG group has significantly more negative asymmetry than the control group in the temporal lobes (Table S4.1, Figure 4), nor did the ETLE group show significantly more negative asymmetry in the frontal, parietal, or occipital lobes (Table S4.1). The only between-group differences in any lobe were that the TLE-HS group had a more negative PVS-VF asymmetry in the Frontal lobes than IGE (Dunn = 3.01, p = 0.027, without age & sex correction) and controls (z = 3.03, p = 0.019, after age and sex correction) (Table S4.1), both with small effect sizes (d = 0.37 and 0.24, respectively).

**Figure 4:**
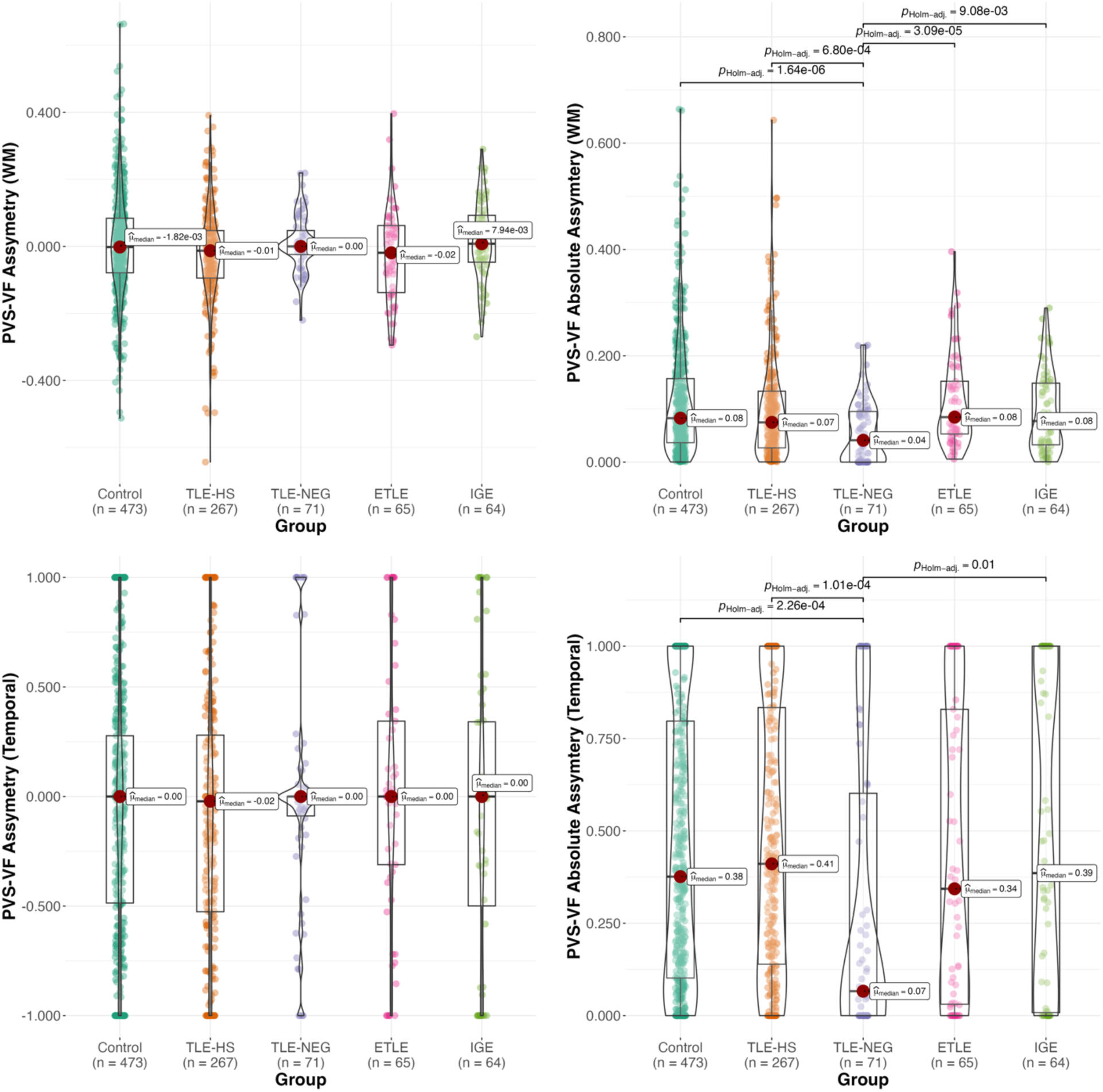
Group comparison of asymmetry (left) and absolute asymmetry (right), in the whole WM (top) and temporal lobes (bottom) Footnote TLE-HS: temporal lobe epilepsy with hippocampal sclerosis; IGE: idiopathic generalised epilepsy; TLE-NEG: temporal lobe epilepsy with MRI negative; ETLE: extratemporal epilepsy.

When ignoring the laterality of the lesion and looking at the absolute asymmetry, the only between-group differences in both the total WM (Table S5.1, Figure 4) and in the various lobes (Table S5.1) was that every group had significantly larger PVS-VF absolute asymmetry than the TLE-NEG group. A comparison of Figure 4’s left and right columns indicates that these differences in absolute asymmetry are driven by reduced variances in the asymmetry (shorter tails) for TLE-NEG, as opposed to any difference in the mean/median asymmetry.

### Association with Duration of Illness

There were no associations between PVS-VF and the duration of illness in the WM (t=-0.65, p=0.518, Table S6), the basal ganglia (t=-0.15,p=0.885, Table S6), subregions (Table S6), raw PVS volumes, or PVS number (data not shown).

## Discussion

In this work, we found that PVS were enlarged in the basal ganglia in all epilepsy types studied. They were not enlarged in the WM after age and sex were accounted for. Further, focal epilepsy localised to the temporal lobes did not show a different distribution of PVS on the ipsilateral side compared to the contralateral temporal lobe.

Although few studies have analysed and quantified PVS in epilepsy subtypes, it is intriguing that PVS dysfunction and epilepsy share some pathological processes, including inflammation, astrogliosis, amyloid (and Tau) deposits, BBB disruption and Aquaporin-4 dysregulation (REFS from Figure 5) (Figure 5).

**Figure 5:**
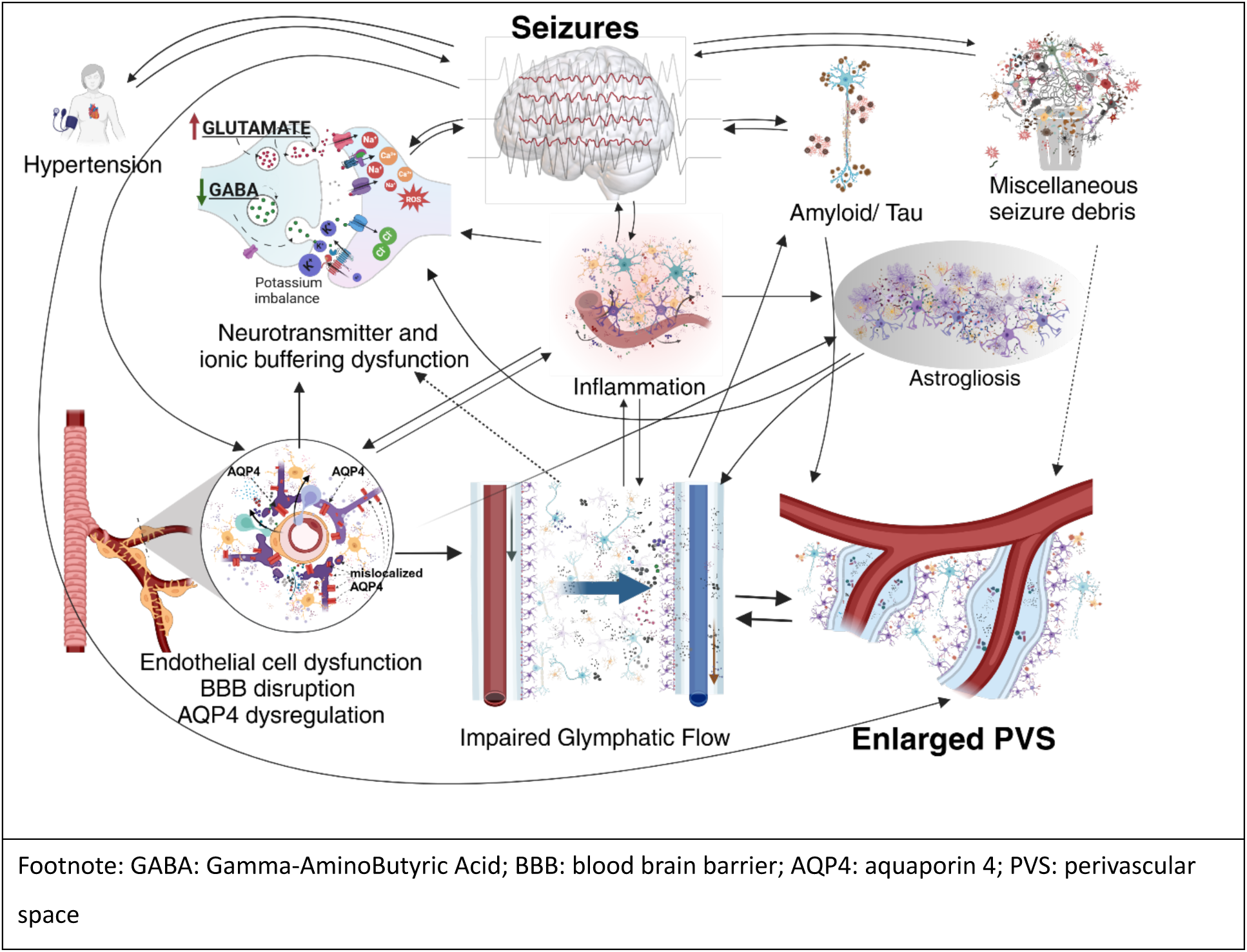
Diagram exploring the possible epilepsy-associated factors related to PVS changes. Filled lines represent relationships described in the literature; dotted lines are hypothesized associations. Adapted from: (Pham et al., 2022) (Akyuz et al., 2021; Christensen et al., 2021; Devinsky et al., 2013; Nowicka, 2023; Rasmussen et al., 2018; Vezzani et al., 2022). Created in: https://BioRender.com

Extra-cellular solutes implicated in seizure initiation include accumulation of excess glutamate (Albrecht and Zielińska, 2017, Eid et al., 2016), potassium (Fisher et al., 1976), inflammatory markers (Ravizza et al., 2024) and aggregated proteins such as amyloid and tau (Tai et al., 2018, Tai et al., 2016, Thom et al., 2011, Chang et al., 2021)(Figure 5). There is no direct evidence that the glymphatic system is involved in potassium and glutamine-glutamate buffering, although astrocytes are directly involved in this process (Henning et al., 2023, Eid et al., 2016). However, given the glymphatic system’s role in maintaining the extracellular environment, it is likely to influence extracellular K+ and glutamine-glutamate levels, as discussed in a review of glymphatic impairment and hepatic encephalopathy (Claeys et al., 2021). Conversely, inflammatory markers such as fibrinogen, C-reactive protein, and interleukin-6 are associated with MRI-visible perivascular space enlargement (Aribisala et al., 2014) and also with contrast-enhancing lesions in MS, implicating inflammatory processes in their enlargement and presumably, removal (Wuerfel et al., 2008). Likewise, amyloid and tau are both known to be cleared partially via the PVS (Eide et al., 2023, Wardlaw et al., 2020a), and amyloid and tau-related diseases often present with enlarged perivascular spaces (Kamagata et al., 2022, Chen et al., 2022). Thus, one possibility for the association between seizures and enlarged PVS is that impaired glymphatic flow causes the accumulation of solutes lowering the excitation threshold, thereby contributing to seizure activity.

Alternatively, seizures themselves may generate by-products that are cleared via the glymphatics. Seizures lead to an excess of leukocytes, cytokines, chemokines (Vezzani et al., 2019), potassium (Henning et al., 2023), glutamate (Eid et al., 2016), lactate, and cell debris from apoptosis (Rho and Boison, 2022). This increased load of solutes may tax the glymphatic system, leading to an enlargement of the PVS.

A third explanation is that seizures and enlarged perivascular spaces are both downstream by-products of other processes. Endothelial cell dysfunction and blood-brain barrier permeability are potential candidates. Blood-brain barrier permeability is associated with epilepsy (Van Vliet et al., 2015, Greene et al., 2022). Whilst BBB conditions are associated with an increase in seizures (and PVS enlargement), not all BBB conditions result in seizures, indicating indirect and variable mechanisms. One potential common cause of both epilepsy and enlarged PVS is astrocyte dysfunction, which has long been described in epilepsy (Binder and Steinhäuser, 2006, Henning et al., 2023). They play a central role in maintaining the neuronal environment, including via potassium (Steinhäuser et al., 2016, Traynelis and Dingledine, 1989) and GABA (Boddum et al., 2016) buffering and glutamate cycling, and play a protective role in the homeostasis of neuronal firing patterns after epileptic activity (Ahtiainen et al., 2021). They also provide the structure of the perivascular spaces, with astrocytic end feet wrapping around the basal lumen for the PVS.

No PVS measure in any region was associated with the duration of the epilepsy (Table S6). If seizures contributed to a gradual degradation of the glymphatic system and associated enlargement of PVS, over the timespan of years, we would expect to see larger PVS with increasing duration of illness. However, with a cross-sectional analysis and long duration of epilepsy (at least 10 years), we cannot exclude the possibility that the onset of spontaneous recurrent seizures produces a relatively fast (and potentially irreversible) process of enlargement of PVS. Longitudinal studies will help us to more accurately characterise the time course of PVS enlargement (with or without progression) to determine whether enlargement was indeed present prior to seizure-onset.

The localisation of PVS enlargement to the Basal Ganglia was unexpected. The Basal Ganglia is supplied with blood by the middle cerebral artery, whereas the WM is predominantly supplied by the anterior and posterior cerebral arteries. In general, abnormalities in the Basal Ganglia PVS are more associated with vascular pathology (Rowsthorn et al., 2023, Smeijer et al., 2019, Tu et al., 2022, Yamasaki et al., 2022), whilst abnormalities in the WM have been more associated with amyloid pathology (Charidimou et al., 2017, Wang et al., 2022). The recent analyses of deep white matter of temporal lobe specimens (44 TLE patients with or without hippocampal sclerosis) showed a relationship between increased perivascular space and preoperative cognitive dysfunction (and reduced fractional anisotropy)(Liu et al., 2024). Enlarged PVS in the Basal Ganglia have been reported in people with vascular disease (Crizzle and Newhouse), multiple sclerosis (Kolbe et al., 2022b), Parkinson disease (Donahue et al., 2024, Foreman et al., 2024, Kim et al., 2024), older community-dwelling adults (Laveskog et al., 2020, Ding et al., 2017) and stroke (Lau et al., 2017). In stroke patients, BG PVS count (but not volume) was negatively associated with von Willebrand factor marker of endothelial function (Wang et al., 2016), implying a link between vascular dysfunction and enlarged PVS in the BG.

The basal ganglia have generally been less implicated as a primary epileptogenic region in epilepsy than cortical regions. However, one recent study with stereoelectroencephalographic (SEEG) recordings from the basal ganglia revealed two patterns of cortico-striatal synchronization in focal seizures (one starting at the seizure onset and the second occurring later); these interactions between the cortex and striatum suggest there is a stronger relationship between the cortex and the BG, apparently involved in the start and cessation of ictal activity (Aupy et al., 2019). Furthermore, the Basal Ganglia is commonly seen to be activated on blood flow ictal SPECT studies injected during seizures (Aupy et al., 2018, Mizobuchi et al., 2004, O’Brien et al., 1998). Besides, the BG is an important modulatory and integrative region, which controls the excitability of premotor neurons, and thalamic nuclei connected to motor, premotor, prefrontal and limbic regions (Slaght et al., 2002). Thus, independent of the thalamus, the BG are thought to play a role in seizure propagation (Vuong and Devergnas, 2018), (Moeller et al., 2008, Aupy et al., 2019). Relevant to our finding of abnormalities across epilepsy syndromes, BG involvement has been observed in many different types of epilepsy including focal, and generalized genetic epilepsies(Gong et al., 2021) (Xiao et al., 2023, Luo et al., 2011, Gong et al., 2021). For example, Bouilleret et al., 2005 (Bouilleret et al., 2005) reported decreased PET dopamine (the primary neurotransmitter in the BG) tracer uptake in the BG in both generalised and TLE. Devergnas et al 2012., (Devergnas et al., 2012) observed increased firing rate in the putamen in both motor and pre-frontal seizures.

From a methodological standpoint, our analysis is better placed to detect differences in the BG-PVS than others due to the use of PINGU. PVS in the BG are more ovoid and less linear than those in the WM, rendering them less amenable to segmentation using vesselness filters, such as the Frangi filter (Frangi et al., 1998), perhaps the most widespread algorithm in use for PVS segmentation. Those AI algorithms that are publicly available to segment PVS were trained on WM-PVS, and thus would not necessarily generalise to BG-PVS. PINGU was trained on both WM- and BG-PVS, and has a much higher performance than the other publicly available algorithms, particularly in the BG (Sinclair et al., 2024).

The BG region in this study included the thalami, since these anatomical regions are traditionally considered together in manual rating scales of enlarged PVS burden (Potter et al., 2015a). The thalamocortical circuits are implicated in the spread of seizures in both generalised epilepsy (Gong et al., 2021) and focal epilepsy(Weng et al., 2020, Aupy et al., 2019). Given a recent report of increased number of enlarged PVS in the thalami in epilepsy (Langan et al., 2024), we performed an analysis in the thalami, and the BG excluding thalami. We found that whilst all epilepsy groups had greater PVS-VF (Table S3.1) and PVS-Number (Table S3.3) in the BG exc. thalami, with large effect sizes (d = 0.94-1.39), only the TLE-HS group had greater PVS-VF (d = 0.26, p = 0.005) in the thalami. Interestingly, the TLE-HS group also showed large effect sizes (d=0.4-0.8) of thalamic atrophy, compared to the IGE-group (d=-0.4) in the Enigma study (Whelan et al., 2018).

Contrary to Feldman et al 2019 (Feldman et al., 2018), we did not find an increased absolute asymmetry in any epilepsy group, but there was a reduced absolute asymmetry in the TLE-NEG group compared to controls, and every other epilepsy group, due to the reduced variance about zero in the TLE group.

The primary limitation of this study is that it is cross-sectional, thus, we cannot infer whether enlarged PVS precede, or follow seizures. Other weaknesses are the significant difference in age between groups, given that age is the strongest risk factor for enlarged PVS. The strengths of this study are the large sample size, 3T MRI, the range of sub-diagnoses of epilepsy available, and our use of a whole-brain PVS segmentation algorithm with higher performance in the basal ganglia than other algorithms. The application of a whole-brain method to quantify the PVS is advantageous as most of the glymphatic system studies in epilepsy have used a different technique, the DTI-ALPS(Taoka et al., 2017) (diffusion tensor imaging along the perivascular space). This method has been criticized due to its limited ability to detect the perivascular spaces(Ringstad, 2024) as it assesses the glymphatic system (indirectly) exclusively in a certain region the white matter (which includes a very small fraction (1%) (Barisano et al., 2021) of total PVS).

In conclusion, this is the first work to demonstrate enlarged PVS in the basal ganglia in epilepsy, and notably, the differences were present in all epilepsy subtypes considered, suggesting either a common mechanism enhancing susceptibility to seizures or a common impact of recurrent seizures on the glymphatic system.

## Data Availability

The raw data used in the present study are not available, due to patient data confidentiality.
Processed data and code are available upon reasonable request

