## Supplementary Material for "Enlarged Perivascular Spaces in the Basal Ganglia Across Epilepsy Subtypes"

SUPPLEMENTAL MATERIAL –

A) Groups: subtypes of epilepsy are

| |
|---|
| TLE-HS: temporal lobe epilepsy with hippocampal sclerosis |
| TLE-NEG: temporal lobe epilepsy with MRI negative |
| IGE: idiopathic generalised epilepsy |
| ETLE: extratemporal epilepsy |

B) Organization of Supplementary tables:

This illustration shows groups of tables S2-S5, according to type of variable and brain regions analysed.
(Created in: https://BioRender.com)

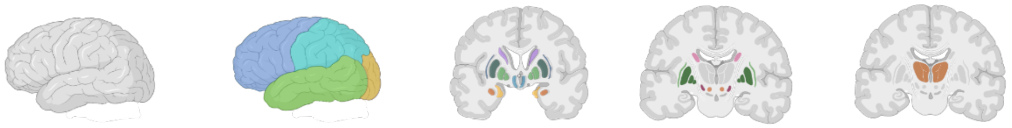

| | Total WM | Regional WM (four lobes ) | Basal Ganglia | BG excluding thalami | Thalami |
|---|---|---|---|---|---|
| **PVS-Volume fraction (VF)** | Table S2.1 | Table S2.1 | Table S3.1 | Table S3.1 | Table S3.1 |
| **PVS-Volume** | Table S2.2 | Table S2.2 | Table S3.2 | Table S3.2 | Table S3.2 |
| **PVS-Number** | Table S2.3 | Table S2.3 | Table S3.3 | Table S3.3 | Table S3.3 |
| **Asymmetry PVS-VF** | Table S4.1 | Table S4.1 | | | |
| **Asymmetry PVS-Volume** | Table S4.2 | Table S4.2 | | | |
| **Asymmetry PVS-Number** | Table S4.3 | Table S4.3 | | | |
| **Absolute Asymmetry PVS-VF** | Table S5.1 | Table S5.1 | | | |
| **Absolute Asymmetry PVS-Volume** | Table S5.2 | Table S5.2 | | | |
| **Absolute Asymmetry PVS-Number** | Table S5.3 | Table S5.3 | | | |

Footnote: PVS= perivascular space; VF= volume fraction; WM= white matter; BG= basal ganglia
Lobes: frontal, occipital, parietal and temporal

| **SUPPLEMENTARY TABLES** | | Page |
|---|---|---|
| Table S1. Difference in demographics variables | | |
| S2. PVS in the White Matter | | |
| | Table S2.1: PVS Volume Fraction in the white matter | |
| | Table S2.2 PVS total volume in the white matter | |
| | Table S2.3: PVS number in the white matter | |
| S3. PVS in the Basal Ganglia | | |
| | Table S3.1: PVS Volume Fraction in the Basal Ganglia | |
| | Table S3.2: PVS total volume in the Basal Ganglia | |
| | Table S3.3: PVS number in the Basal Ganglia | |
| S4. Asymmetry | | |
| | Table S4.0.1: Median and mean asymmetry measures for each group. Measures where the mean is significantly different from zero are highlighted in bold. | |
| | Table S4.1: PVS Volume Fraction asymmetry in the white matter | |
| | Table S4.2: PVS-Volume asymmetry in the white matter | |
| | Table S4.3: PVS number asymmetry in the white matter | |
| S5. Absolute asymmetry | | |
| | Table S5.1: PVS Volume Fraction absolute asymmetry in the white matter | |
| | Table S5.2: PVS total Volume absolute asymmetry in the white matter | |
| | Table S5.3: PVS number absolute asymmetry in the white matter | |
| Table S6: Association of duration of illness with PVS Volume Fraction in each region and sub-region, corrected for age and sex. | | |

| **SUPPLEMENTARY FIGURES** | Page |
|---|---|
| Figure S1: Histograms of number of PVS in each White Matter (WM) region | |
| Figure S2: Histograms of number of PVS in each Basal Ganglia (BG) region | |
| Figure S3: Histograms PVS-Number asymmetry in each White Matter (WM) region | |
| Figure S4: PVS volume fraction in Basal Ganglia excluding Thalamus (BG exc. Thalamus, left) and Thalamus (Thal, right) by group | |

Table S1: Difference in demographics variables.

| Group | | Age | | | | Duration | | | | Sex | | | |
|---|---|---|---|---|---|---|---|---|---|---|---|---|---|
| | | median (IQR) | | Univariate | | median (IQR) | | Univariate | | % Female | | Univariate | |
| G1 | G2 | G1 | G2 | U | p | G1 | G2 | U | p | G1 | G2 | $X^2$ | p |
| Control | TLE-HS | 35.5(24) | 46(12) | 46242 | **1.41x10-9** | - | - | - | - | 38.3 | 38.6 | 2.19x10-5 | 0.996 |
| Control | TLE-NEG | 35.5(24) | 46(18.5) | 12633 | **7.58x10-4** | - | - | - | - | 38.3 | 39.4 | 0.003 | 0.954 |
| Control | ETLE | 35.5(24) | 31(16) | 20093 | **5.87x10-5** | - | - | - | - | 38.3 | 60.0 | 10.29 | **0.001** |
| Control | IGE | 35.5(24) | 33(13) | 17900 | **0.018** | - | - | - | - | 38.3 | 37.5 | 2.76x10-30 | 1.000 |
| TLE-HS | TLE- NEG | 46(12) | 46(18.5) | 9316 | 0.825 | 35(19) | 24(21) | 12420 | **5.80x10-5** | 38.6 | 39.4 | 1.24x10-30 | 1.000 |
| TLE-HS | ETLE | 46(12) | 31(16) | 14571 | **1.97x10-17** | 35(19) | 20(17) | 13187 | **8.04x10-11** | 38.6 | 60.0 | 8.95 | **0.003** |
| TLE-HS | IGE | 46(12) | 33(13) | 13462 | **8.35x10-13** | 35(19) | 21(15.2) | 12862 | **3.34x10-10** | 38.6 | 37.5 | 2.56x10-4 | 0.987 |
| TLE- NEG | ETLE | 46(18.5) | 31(16) | 3692 | **1.64x10-9** | 24(21) | 20(17) | 2837 | **0.021** | 39.4 | 60.0 | 4.95 | **0.026** |
| TLE- NEG | IGE | 46(18.5) | 33(13) | 3423 | **3.99x10-7** | 24(21) | 21(15.2) | 2658 | **0.09** | 39.4 | 37.5 | 0.003 | 0.957 |
| ETLE | IGE | 31(16) | 33(13) | 1763 | 0.136 | 20(17) | 21(15.2) | 1930 | 0.481 | 60.0 | 37.5 | 5.66 | **0.017** |

Footnote: TLE: temporal lobe epilepsy; HS: hippocampal sclerosis; TLE-NEG: temporal lobe epilepsy with MRI negative; ETLE: extratemporal epilepsy; IGE: Idiopathic generalised epilepsy

Table S2.1: PVS Volume Fraction in the white matter (WM)

| | Group | | median (IQR) (x$10^{-2}$) | | | Univariate | | Generalised linear model | | | |
|---|---|---|---|---|---|---|---|---|---|---|---|
| | G1 | G2 | G1 | G2 | Δ% | Dunn | p-Holm | beta(sderr) | d(G) | z | p-Tukey |
| WM | Control | TLE-HS | 0.182(0.182) | 0.199(0.150) | 9.6 | 2.26 | 0.168 | -4.45(22.80)x$10^{+0}$ | -0.032 | -0.20 | 1.000 |
| | Control | TLE-NEG | 0.182(0.182) | 0.210(0.172) | 15.5 | 1.60 | 0.442 | 4.11(37.20)x$10^{+0}$ | -0.039 | 0.11 | 1.000 |
| | Control | EXTRA | 0.182(0.182) | 0.159(0.133) | -12.4 | 1.31 | 0.570 | 1.43(4.85)x$10^{+1}$ | -0.033 | 0.30 | 0.998 |
| | Control | IGE | 0.182(0.182) | 0.135(0.130) | -25.6 | 2.17 | 0.170 | 3.86(4.95)x$10^{+1}$ | -0.109 | 0.78 | 0.930 |
| | TLE-HS | TLE- NEG | 0.199(0.150) | 0.210(0.172) | 5.4 | 0.23 | 1.000 | 8.56(38.80)x$10^{+0}$ | -0.006 | 0.22 | 0.999 |
| | TLE-HS | EXTRA | 0.199(0.150) | 0.159(0.133) | -20.0 | 2.50 | 0.099 | 1.88(4.99)x$10^{+1}$ | -0.000 | 0.38 | 0.995 |
| | TLE-HS | IGE | 0.199(0.150) | 0.135(0.130) | -32.1 | 3.32 | **0.009** | 4.30(5.08)x$10^{+1}$ | -0.077 | 0.85 | 0.907 |
| | TLE- NEG | EXTRA | 0.210(0.172) | 0.159(0.133) | -24.1 | 2.19 | 0.170 | 1.02(5.82)x$10^{+1}$ | 0.006 | 0.18 | 1.000 |
| | TLE- NEG | IGE | 0.210(0.172) | 0.135(0.130) | -35.6 | 2.86 | **0.039** | 3.44(5.90)x$10^{+1}$ | -0.071 | 0.58 | 0.975 |
| | EXTRA | IGE | 0.159(0.133) | 0.135(0.130) | -15.1 | 0.66 | 1.000 | 2.42(6.53)x$10^{+1}$ | -0.077 | 0.37 | 0.995 |
| WM - Frontal | Control | TLE-HS | 0.219(0.247) | 0.239(0.200) | 9.2 | 2.29 | 0.109 | -1.28(2.08)x$10^{+1}$ | -0.010 | -0.61 | 0.970 |
| | Control | TLE- NEG | 0.219(0.247) | 0.261(0.284) | 19.0 | 2.36 | 0.109 | -1.50(3.26)x$10^{+1}$ | 0.036 | -0.46 | 0.989 |
| | Control | EXTRA | 0.219(0.247) | 0.180(0.192) | -18.0 | 1.73 | 0.252 | 4.73(5.00)x$10^{+1}$ | -0.102 | 0.95 | 0.865 |
| | Control | IGE | 0.219(0.247) | 0.148(0.197) | -32.2 | 2.00 | 0.181 | 3.55(4.79)x$10^{+1}$ | -0.102 | 0.74 | 0.940 |
| | TLE-HS | TLE- NEG | 0.239(0.200) | 0.261(0.284) | 9.0 | 0.94 | 0.699 | -2.25(33.90)x$10^{+0}$ | 0.046 | -0.07 | 1.000 |
| | TLE-HS | EXTRA | 0.239(0.200) | 0.180(0.192) | -24.9 | 2.92 | **0.024** | 6.01(5.10)x$10^{+1}$ | -0.092 | 1.18 | 0.742 |
| | TLE-HS | IGE | 0.239(0.200) | 0.148(0.197) | -37.9 | 3.18 | **0.013** | 4.82(4.89)x$10^{+1}$ | -0.092 | 0.99 | 0.847 |
| | TLE- NEG | EXTRA | 0.261(0.284) | 0.180(0.192) | -31.1 | 3.08 | **0.016** | 6.23(5.71)x$10^{+1}$ | -0.138 | 1.09 | 0.793 |
| | TLE- NEG | IGE | 0.261(0.284) | 0.148(0.197) | -43.0 | 3.29 | **0.010** | 5.05(5.52)x$10^{+1}$ | -0.138 | 0.91 | 0.880 |
| | EXTRA | IGE | 0.180(0.192) | 0.148(0.197) | -17.3 | 0.22 | 0.829 | -1.19(6.57)x$10^{+1}$ | -0.000 | -0.18 | 1.000 |
| WM - Parietal | Control | TLE-HS | 0.318(0.329) | 0.323(0.302) | 1.6 | 0.88 | 1.000 | 1.17(1.49)x$10^{+1}$ | -0.092 | 0.78 | 0.928 |
| | Control | TLE- NEG | 0.318(0.329) | 0.306(0.286) | -3.7 | 0.30 | 1.000 | 2.08(2.52)x$10^{+1}$ | -0.128 | 0.83 | 0.915 |
| | Control | EXTRA | 0.318(0.329) | 0.291(0.253) | -8.3 | 0.74 | 1.000 | -7.57(28.10)x$10^{+0}$ | 0.030 | -0.27 | 0.999 |
| | Control | IGE | 0.318(0.329) | 0.220(0.260) | -30.7 | 2.32 | 0.185 | 3.96(3.19)x$10^{+1}$ | -0.169 | 1.24 | 0.705 |
| | TLE-HS | TLE- NEG | 0.323(0.302) | 0.306(0.286) | -5.3 | 0.22 | 1.000 | 9.13(26.40)x$10^{+0}$ | -0.035 | 0.35 | 0.997 |
| | TLE-HS | EXTRA | 0.323(0.302) | 0.291(0.253) | -9.7 | 1.20 | 1.000 | -1.92(2.94)x$10^{+1}$ | 0.122 | -0.65 | 0.962 |
| | TLE-HS | IGE | 0.323(0.302) | 0.220(0.260) | -31.8 | 2.70 | 0.069 | 2.79(3.30)x$10^{+1}$ | -0.077 | 0.85 | 0.907 |
| | TLE- NEG | EXTRA | 0.306(0.286) | 0.291(0.253) | -4.7 | 0.80 | 1.000 | -2.84(3.59)x$10^{+1}$ | 0.157 | -0.79 | 0.926 |
| | TLE- NEG | IGE | 0.306(0.286) | 0.220(0.260) | -28.0 | 2.01 | 0.352 | 1.88(3.89)x$10^{+1}$ | -0.042 | 0.48 | 0.987 |
| | EXTRA | IGE | 0.291(0.253) | 0.220(0.260) | -24.4 | 1.19 | 1.000 | 4.72(4.00)x$10^{+1}$ | -0.199 | 1.18 | 0.744 |
| WM - Temporal | Control | TLE-HS | 0.024(0.066) | 0.027(0.056) | 8.9 | 0.66 | 1.000 | 1.16(1.87)x$10^{+2}$ | -0.133 | 0.62 | 0.967 |
| | Control | TLE- NEG | 0.024(0.066) | 0.034(0.080) | 40.3 | 1.05 | 1.000 | 1.03(2.94)x$10^{+2}$ | -0.099 | 0.35 | 0.996 |
| | Control | EXTRA | 0.024(0.066) | 0.021(0.042) | -15.6 | 1.91 | 0.281 | 5.21(5.30)x$10^{+2}$ | -0.099 | 0.98 | 0.844 |
| | Control | IGE | 0.024(0.066) | 0.013(0.035) | -45.6 | 2.89 | **0.031** | 8.82(5.80)x$10^{+2}$ | -0.191 | 1.52 | 0.512 |
| | TLE-HS | TLE- NEG | 0.027(0.056) | 0.034(0.080) | 28.8 | 0.63 | 1.000 | -1.35(31.60)x$10^{+1}$ | 0.035 | -0.04 | 1.000 |
| | TLE-HS | EXTRA | 0.027(0.056) | 0.021(0.042) | -22.5 | 2.19 | 0.170 | 4.05(5.38)x$10^{+2}$ | 0.035 | 0.75 | 0.935 |
| | TLE-HS | IGE | 0.027(0.056) | 0.013(0.035) | -50.1 | 3.13 | **0.017** | 7.66(5.88)x$10^{+2}$ | -0.058 | 1.30 | 0.657 |
| | TLE- NEG | EXTRA | 0.034(0.080) | 0.021(0.042) | -39.8 | 2.25 | 0.170 | 4.18(5.87)x$10^{+2}$ | 0.000 | 0.71 | 0.946 |
| | TLE- NEG | IGE | 0.034(0.080) | 0.013(0.035) | -61.3 | 3.01 | **0.023** | 7.79(6.33)x$10^{+2}$ | -0.092 | 1.23 | 0.703 |
| | EXTRA | IGE | 0.021(0.042) | 0.013(0.035) | -35.6 | 0.75 | 1.000 | 3.61(7.59)x$10^{+2}$ | -0.093 | 0.48 | 0.988 |
| WM - Occipital | Control | TLE-HS | 0.024(0.096) | 0.003(0.017) | -89.6 | 9.74 | **<1x$10^{-20}$** | 46.40(7.40)x$10^{+2}$ | -0.689 | 6.27 | **1.91x$10^{-9}$** |
| | Control | TLE- NEG | 0.024(0.096) | 0.002(0.016) | -91.7 | 6.16 | **6.03x$10^{-9}$** | 7.41(2.03)x$10^{+3}$ | -0.761 | 3.64 | **0.002** |
| | Control | EXTRA | 0.024(0.096) | 0.002(0.019) | -90.7 | 5.31 | **7.63x$10^{-7}$** | 3.95(1.32)x$10^{+3}$ | -0.661 | 3.00 | **0.019** |
| | Control | IGE | 0.024(0.096) | 0.001(0.016) | -96.0 | 6.17 | **6.03x$10^{-9}$** | 8.16(2.31)x$10^{+3}$ | -0.784 | 3.53 | **0.003** |
| | TLE-HS | TLE- NEG | 0.003(0.017) | 0.002(0.016) | -20.7 | 0.29 | 1.000 | 2.77(2.15)x$10^{+3}$ | -0.072 | 1.28 | 0.663 |
| | TLE-HS | EXTRA | 0.003(0.017) | 0.002(0.019) | -11.0 | 0.31 | 1.000 | -6.88(15.00)x$10^{+2}$ | 0.027 | -0.46 | 0.989 |
| | TLE-HS | IGE | 0.003(0.017) | 0.001(0.016) | -61.9 | 0.55 | 1.000 | 3.52(2.42)x$10^{+3}$ | -0.096 | 1.45 | 0.550 |
| | TLE- NEG | EXTRA | 0.002(0.016) | 0.002(0.019) | 12.1 | 0.48 | 1.000 | -3.46(2.42)x$10^{+3}$ | 0.099 | -1.43 | 0.566 |
| | TLE- NEG | IGE | 0.002(0.016) | 0.001(0.016) | -52.0 | 0.22 | 1.000 | 7.52(30.70)x$10^{+2}$ | -0.023 | 0.24 | 0.999 |
| | EXTRA | IGE | 0.002(0.019) | 0.001(0.016) | -57.2 | 0.68 | 1.000 | 4.21(2.65)x$10^{+3}$ | -0.123 | 1.59 | 0.463 |

Table S2.2 PVS total volume in the white matter (WM)

| | Group | | median (IQR) (mm$^3$) | | Univariate | | | Generalised Linear Model | | | |
|---|---|---|---|---|---|---|---|---|---|---|---|
| | G1 | G2 | G1 | G2 | $\Delta$% | Dunn | p-Holm | beta(sderr) | d(G) | z | p-Tukey |
| WM | Control | TLE-HS | 779(723) | 784(672) | 0.6 | 0.81 | 1.000 | 4.94(5.85)x10$^{-5}$ | -0.106 | 0.84 | 0.908 |
| | Control | TLE- NEG | 779(723) | 780(804) | 0.1 | 0.89 | 1.000 | 3.81(9.47)x10$^{-5}$ | -0.074 | 0.40 | 0.994 |
| | Control | ETLE | 779(723) | 660(556) | -15.3 | 1.50 | 0.665 | 4.87(11.70)x10$^{-5}$ | -0.064 | 0.42 | 0.993 |
| | Control | IGE | 779(723) | 534(640) | -31.4 | 2.58 | 0.080 | 1.14(1.23)x10$^{-4}$ | -0.134 | 0.92 | 0.877 |
| | TLE-HS | TLE- NEG | 784(672) | 780(804) | -0.5 | 0.38 | 1.000 | -1.13(9.97)x10$^{-5}$ | 0.033 | -0.11 | 1.000 |
| | TLE-HS | ETLE | 784(672) | 660(556) | -15.8 | 1.89 | 0.415 | -6.50(1210.00)x10$^{-7}$ | 0.042 | -0.01 | 1.000 |
| | TLE-HS | IGE | 784(672) | 534(640) | -31.8 | 2.91 | **0.036** | 6.44(12.70)x10$^{-5}$ | -0.028 | 0.51 | 0.985 |
| | TLE- NEG | ETLE | 780(804) | 660(556) | -15.4 | 1.82 | 0.417 | 1.06(14.30)x10$^{-5}$ | 0.009 | 0.07 | 1.000 |
| | TLE- NEG | IGE | 780(804) | 534(640) | -31.5 | 2.65 | 0.073 | 7.57(14.80)x10$^{-5}$ | -0.060 | 0.51 | 0.985 |
| | ETLE | IGE | 660(556) | 534(640) | -19.0 | 0.82 | 1.000 | 6.51(16.00)x10$^{-5}$ | -0.070 | 0.41 | 0.994 |
| WM - Frontal | Control | TLE-HS | 306(365) | 322(312) | 5.2 | 1.25 | 0.635 | 2.21(15.60)x10$^{-5}$ | -0.062 | 0.14 | 1.000 |
| | Control | TLE- NEG | 306(365) | 339(428) | 10.8 | 1.80 | 0.357 | -3.99(24.50)x10$^{-5}$ | 0.000 | -0.16 | 1.000 |
| | Control | ETLE | 306(365) | 256(262) | -16.3 | 1.77 | 0.357 | 3.40(3.54)x10$^{-4}$ | -0.125 | 0.96 | 0.859 |
| | Control | IGE | 306(365) | 205(265) | -33.0 | 2.31 | 0.124 | 2.77(3.48)x10$^{-4}$ | -0.115 | 0.80 | 0.924 |
| | TLE-HS | TLE- NEG | 322(312) | 339(428) | 5.3 | 1.00 | 0.635 | -6.20(25.70)x10$^{-5}$ | 0.062 | -0.24 | 0.999 |
| | TLE-HS | ETLE | 322(312) | 256(262) | -20.5 | 2.39 | 0.119 | 3.18(3.63)x10$^{-4}$ | -0.063 | 0.88 | 0.895 |
| | TLE-HS | IGE | 322(312) | 205(265) | -36.3 | 2.90 | **0.033** | 2.55(3.58)x10$^{-4}$ | -0.053 | 0.71 | 0.948 |
| | TLE- NEG | ETLE | 339(428) | 256(262) | -24.5 | 2.70 | 0.055 | 3.80(4.11)x10$^{-4}$ | -0.125 | 0.92 | 0.876 |
| | TLE- NEG | IGE | 339(428) | 205(265) | -39.5 | 3.12 | **0.018** | 3.17(4.06)x10$^{-4}$ | -0.115 | 0.78 | 0.929 |
| | ETLE | IGE | 256(262) | 205(265) | -19.9 | 0.42 | 0.676 | -6.30(47.00)x10$^{-5}$ | 0.010 | -0.13 | 1.000 |
| WM - Parietal | Control | TLE-HS | 289(296) | 280(286) | -3.1 | 0.09 | 1.000 | 2.59(1.74)x10$^{-4}$ | -0.144 | 1.48 | 0.549 |
| | Control | TLE- NEG | 289(296) | 271(284) | -6.2 | 0.19 | 1.000 | 2.99(2.93)x10$^{-4}$ | -0.153 | 1.02 | 0.832 |
| | Control | ETLE | 289(296) | 248(282) | -14.2 | 0.99 | 1.000 | -4.44(31.20)x10$^{-5}$ | 0.009 | -0.14 | 1.000 |
| | Control | IGE | 289(296) | 188(255) | -34.8 | 2.65 | 0.081 | 4.63(3.63)x10$^{-4}$ | -0.179 | 1.28 | 0.686 |
| | TLE-HS | TLE- NEG | 280(286) | 271(284) | -3.2 | 0.13 | 1.000 | 4.04(30.90)x10$^{-5}$ | -0.009 | 0.13 | 1.000 |
| | TLE-HS | ETLE | 280(286) | 248(282) | -11.4 | 0.90 | 1.000 | -3.03(3.30)x10$^{-4}$ | 0.152 | -0.92 | 0.879 |
| | TLE-HS | IGE | 280(286) | 188(255) | -32.7 | 2.49 | 0.116 | 2.04(3.78)x10$^{-4}$ | -0.035 | 0.54 | 0.981 |
| | TLE- NEG | ETLE | 271(284) | 248(282) | -8.5 | 0.62 | 1.000 | -3.43(4.07)x10$^{-4}$ | 0.162 | -0.84 | 0.908 |
| | TLE- NEG | IGE | 271(284) | 188(255) | -30.4 | 1.91 | 0.454 | 1.63(4.46)x10$^{-4}$ | -0.026 | 0.37 | 0.996 |
| | ETLE | IGE | 248(282) | 188(255) | -24.0 | 1.26 | 1.000 | 5.07(4.50)x10$^{-4}$ | -0.187 | 1.13 | 0.776 |
| WM - Temporal | Control | TLE-HS | 15(41) | 15(32) | 0.0 | 0.13 | 1.000 | 3.08(3.21)x10$^{-3}$ | -0.159 | 0.96 | 0.856 |
| | Control | TLE- NEG | 15(41) | 20(42) | 33.3 | 0.87 | 1.000 | 1.32(4.78)x10$^{-3}$ | -0.086 | 0.28 | 0.999 |
| | Control | ETLE | 15(41) | 12(26) | -20.0 | 1.87 | 0.370 | 5.55(7.95)x10$^{-3}$ | -0.085 | 0.70 | 0.950 |
| | Control | IGE | 15(41) | 8(21) | -46.7 | 2.95 | 0.031 | 14.40(9.49)x10$^{-3}$ | -0.196 | 1.51 | 0.519 |
| | TLE-HS | TLE- NEG | 15(32) | 20(42) | 33.3 | 0.75 | 1.000 | -1.76(5.23)x10$^{-3}$ | 0.073 | -0.34 | 0.997 |
| | TLE-HS | ETLE | 15(32) | 12(26) | -20.0 | 1.86 | 0.370 | 2.47(8.14)x10$^{-3}$ | 0.074 | 0.30 | 0.998 |
| | TLE-HS | IGE | 15(32) | 8(21) | -46.7 | 2.90 | **0.031** | 11.30(9.65)x10$^{-3}$ | -0.037 | 1.17 | 0.743 |
| | TLE- NEG | ETLE | 20(42) | 12(26) | -40.0 | 2.08 | 0.260 | 4.23(8.95)x10$^{-3}$ | 0.001 | 0.47 | 0.988 |
| | TLE- NEG | IGE | 20(42) | 8(21) | -60.0 | 2.92 | 0.031 | 1.30(1.03)x10$^{-2}$ | -0.110 | 1.26 | 0.686 |
| | ETLE | IGE | 12(26) | 8(21) | -33.3 | 0.83 | 1.000 | 8.81(11.90)x10$^{-3}$ | -0.111 | 0.74 | 0.940 |
| WM - Occipital | Control | TLE-HS | 10(38) | 1(7) | -90.0 | 9.88 | **<1x10$^{-20}$** | 11.90(1.92)x10$^{-2}$ | -0.689 | 6.20 | **2.37x10$^{-9}$** |
| | Control | TLE- NEG | 10(38) | 1(6) | -90.0 | 6.19 | **5.37x10$^{-9}$** | 17.90(5.06)x10$^{-2}$ | -0.751 | 3.54 | **0.003** |
| | Control | ETLE | 10(38) | 1(8) | -90.0 | 5.24 | **1.15x10$^{-6}$** | 8.82(3.07)x10$^{-2}$ | -0.653 | 2.88 | **0.027** |
| | Control | IGE | 10(38) | 0(7) | -95.0 | 6.15 | **6.34x10$^{-9}$** | 19.90(5.78)x10$^{-2}$ | -0.776 | 3.44 | **0.004** |
| | TLE-HS | TLE- NEG | 1(7) | 1(6) | 0.0 | 0.24 | 1.000 | 6.01(5.39)x10$^{-2}$ | -0.062 | 1.12 | 0.768 |
| | TLE-HS | ETLE | 1(7) | 1(8) | 0.0 | 0.46 | 1.000 | -3.09(3.60)x10$^{-2}$ | 0.036 | -0.86 | 0.896 |
| | TLE-HS | IGE | 1(7) | 0(7) | -50.0 | 0.45 | 1.000 | 7.95(6.07)x10$^{-2}$ | -0.087 | 1.31 | 0.647 |
| | TLE- NEG | ETLE | 1(6) | 1(8) | 0.0 | 0.56 | 1.000 | -9.10(5.90)x10$^{-2}$ | 0.098 | -1.54 | 0.492 |
| | TLE- NEG | IGE | 1(6) | 0(7) | -50.0 | 0.18 | 1.000 | 1.94(7.67)x10$^{-2}$ | -0.025 | 0.25 | 0.999 |
| | ETLE | IGE | 1(8) | 0(7) | -50.0 | 0.72 | 1.000 | 11.00(6.52)x10$^{-2}$ | -0.124 | 1.69 | 0.396 |

Figure S2.3: PVS number in the white matter (WM)

| | Group | | median (IQR) | | Univariate | | | General Linear Model | | | |
|---|---|---|---|---|---|---|---|---|---|---|---|
| | G1 | G2 | G1 | G2 | Δ% | Dunn | p-Holm | beta(sderr) | d(G) | z | p-Tukey |
| WM | Control | TLE-HS | 122.0(96.0) | 125.0(90.0) | 2.5 | 1.75 | 0.403 | 4.29(29.00)x10$^{-5}$ | -0.074 | 0.15 | 1.000 |
| | Control | TLE- NEG | 122.0(96.0) | 128.0(97.5) | 4.9 | 1.34 | 0.543 | 1.81(4.76)x10$^{-4}$ | -0.083 | 0.38 | 0.995 |
| | Control | ETLE | 122.0(96.0) | 102.0(68.0) | -16.4 | 1.57 | 0.463 | 3.43(6.08)x10$^{-4}$ | -0.078 | 0.57 | 0.978 |
| | Control | IGE | 122.0(96.0) | 90.5(75.5) | -25.8 | 2.29 | 0.156 | 4.61(6.15)x10$^{-4}$ | -0.115 | 0.75 | 0.938 |
| | TLE-HS | TLE- NEG | 125.0(90.0) | 128.0(97.5) | 2.4 | 0.27 | 1.000 | 1.38(4.97)x10$^{-4}$ | -0.009 | 0.28 | 0.999 |
| | TLE-HS | ETLE | 125.0(90.0) | 102.0(68.0) | -18.4 | 2.47 | 0.108 | 3.01(6.27)x10$^{-4}$ | -0.004 | 0.48 | 0.988 |
| | TLE-HS | IGE | 125.0(90.0) | 90.5(75.5) | -27.6 | 3.15 | **0.016** | 4.18(6.34)x10$^{-4}$ | -0.040 | 0.66 | 0.961 |
| | TLE- NEG | ETLE | 128.0(97.5) | 102.0(68.0) | -20.3 | 2.20 | 0.165 | 1.63(7.35)x10$^{-4}$ | 0.005 | 0.22 | 0.999 |
| | TLE- NEG | IGE | 128.0(97.5) | 90.5(75.5) | -29.3 | 2.75 | 0.053 | 2.80(7.41)x10$^{-4}$ | -0.031 | 0.38 | 0.995 |
| | ETLE | IGE | 102.0(68.0) | 90.5(75.5) | -11.3 | 0.55 | 1.000 | 1.18(8.14)x10$^{-4}$ | -0.036 | 0.14 | 1.000 |
| WM - Frontal | Control | TLE-HS | 55.0(50.0) | 56.0(43.5) | 1.8 | 2.11 | 0.176 | -3.93(7.12)x10$^{-4}$ | -0.023 | -0.55 | 0.979 |
| | Control | TLE- NEG | 55.0(50.0) | 63.0(43.5) | 14.5 | 1.97 | 0.197 | -1.47(11.50)x10$^{-4}$ | -0.016 | -0.13 | 1.000 |
| | Control | ETLE | 55.0(50.0) | 44.0(33.0) | -20.0 | 1.88 | 0.197 | 1.53(1.61)x10$^{-3}$ | -0.118 | 0.95 | 0.865 |
| | Control | IGE | 55.0(50.0) | 40.0(40.5) | -27.3 | 2.24 | 0.152 | 1.22(1.58)x10$^{-3}$ | -0.113 | 0.77 | 0.932 |
| | TLE-HS | TLE- NEG | 56.0(43.5) | 63.0(43.5) | 12.5 | 0.67 | 1.000 | 2.46(12.00)x10$^{-4}$ | 0.007 | 0.21 | 1.000 |
| | TLE-HS | ETLE | 56.0(43.5) | 44.0(33.0) | -21.4 | 2.96 | **0.024** | 1.92(1.65)x10$^{-3}$ | -0.094 | 1.17 | 0.751 |
| | TLE-HS | IGE | 56.0(43.5) | 40.0(40.5) | -28.6 | 3.30 | **0.010** | 1.61(1.62)x10$^{-3}$ | -0.090 | 0.99 | 0.844 |
| | TLE- NEG | ETLE | 63.0(43.5) | 44.0(33.0) | -30.2 | 2.91 | **0.026** | 1.67(1.89)x10$^{-3}$ | -0.101 | 0.89 | 0.891 |
| | TLE- NEG | IGE | 63.0(43.5) | 40.0(40.5) | -36.5 | 3.18 | **0.013** | 1.36(1.86)x10$^{-3}$ | -0.097 | 0.73 | 0.943 |
| | ETLE | IGE | 44.0(33.0) | 40.0(40.5) | -9.1 | 0.28 | 1.000 | -3.10(21.30)x10$^{-4}$ | 0.004 | -0.15 | 1.000 |
| WM - Parietal | Control | TLE-HS | 46.0(36.0) | 48.0(36.5) | 4.3 | 0.60 | 1.000 | 8.32(8.10)x10$^{-4}$ | -0.122 | 1.03 | 0.829 |
| | Control | TLE- NEG | 46.0(36.0) | 44.0(34.5) | -4.3 | 0.30 | 1.000 | 1.54(1.37)x10$^{-3}$ | -0.174 | 1.12 | 0.777 |
| | Control | ETLE | 46.0(36.0) | 42.0(31.0) | -8.7 | 0.81 | 1.000 | 9.93(154.00)x10$^{-5}$ | -0.022 | 0.06 | 1.000 |
| | Control | IGE | 46.0(36.0) | 36.5(33.2) | -20.7 | 2.34 | 0.174 | 1.87(1.67)x10$^{-3}$ | -0.161 | 1.12 | 0.781 |
| | TLE-HS | TLE- NEG | 48.0(36.5) | 44.0(34.5) | -8.3 | 0.06 | 1.000 | 7.09(14.30)x10$^{-4}$ | -0.052 | 0.49 | 0.987 |
| | TLE-HS | ETLE | 48.0(36.5) | 42.0(31.0) | -12.5 | 1.10 | 1.000 | -7.32(16.10)x10$^{-4}$ | 0.101 | -0.45 | 0.990 |
| | TLE-HS | IGE | 48.0(36.5) | 36.5(33.2) | -24.0 | 2.57 | 0.102 | 1.03(1.73)x10$^{-3}$ | -0.039 | 0.60 | 0.973 |
| | TLE- NEG | ETLE | 44.0(34.5) | 42.0(31.0) | -4.5 | 0.84 | 1.000 | -1.44(1.96)x10$^{-3}$ | 0.152 | -0.73 | 0.943 |
| | TLE- NEG | IGE | 44.0(34.5) | 36.5(33.2) | -17.0 | 2.03 | 0.340 | 3.25(20.60)x10$^{-4}$ | 0.013 | 0.16 | 1.000 |
| | ETLE | IGE | 42.0(31.0) | 36.5(33.2) | -13.1 | 1.16 | 1.000 | 1.77(2.13)x10$^{-3}$ | -0.139 | 0.83 | 0.914 |
| WM - Temporal | Control | TLE-HS | 4.0(9.0) | 4.0(8.0) | 0.0 | 0.15 | 1.000 | 1.10(1.18)x10$^{-2}$ | -0.194 | 0.93 | 0.870 |
| | Control | TLE- NEG | 4.0(9.0) | 5.0(8.0) | 25.0 | 0.76 | 1.000 | 9.75(18.50)x10$^{-3}$ | -0.151 | 0.53 | 0.982 |
| | Control | ETLE | 4.0(9.0) | 3.0(5.0) | -25.0 | 2.27 | 0.147 | 3.75(3.26)x10$^{-2}$ | -0.131 | 1.15 | 0.752 |
| | Control | IGE | 4.0(9.0) | 2.0(5.2) | -50.0 | 3.31 | **0.009** | 7.52(3.78)x10$^{-2}$ | -0.252 | 1.99 | 0.241 |
| | TLE-HS | TLE- NEG | 4.0(8.0) | 5.0(8.0) | 25.0 | 0.64 | 1.000 | -1.23(20.10)x10$^{-3}$ | 0.044 | -0.06 | 1.000 |
| | TLE-HS | ETLE | 4.0(8.0) | 3.0(5.0) | -25.0 | 2.25 | 0.147 | 2.65(3.31)x10$^{-2}$ | 0.064 | 0.80 | 0.920 |
| | TLE-HS | IGE | 4.0(8.0) | 2.0(5.2) | -50.0 | 3.25 | **0.010** | 6.42(3.83)x10$^{-2}$ | -0.058 | 1.68 | 0.411 |
| | TLE- NEG | ETLE | 5.0(8.0) | 3.0(5.0) | -40.0 | 2.31 | 0.147 | 2.78(3.63)x10$^{-2}$ | 0.020 | 0.77 | 0.931 |
| | TLE- NEG | IGE | 5.0(8.0) | 2.0(5.2) | -60.0 | 3.12 | **0.015** | 6.54(4.11)x10$^{-2}$ | -0.102 | 1.59 | 0.463 |
| | ETLE | IGE | 3.0(5.0) | 2.0(5.2) | -33.3 | 0.80 | 1.000 | 3.77(4.82)x10$^{-2}$ | -0.122 | 0.78 | 0.926 |
| WM - Occipital | Control | TLE-HS | 2.0(4.0) | 1.0(2.0) | -50.0 | 8.80 | **1.40x10$^{-17}$** | 38.90(6.36)x10$^{-2}$ | -0.567 | 6.11 | **3.87x10$^{-9}$** |
| | Control | TLE- NEG | 2.0(4.0) | 1.0(2.0) | -50.0 | 5.42 | **4.79x10$^{-7}$** | 4.36(1.27)x10$^{-1}$ | -0.595 | 3.43 | **0.005** |
| | Control | ETLE | 2.0(4.0) | 1.0(2.0) | -50.0 | 5.08 | **2.66x10$^{-6}$** | 2.42(1.07)x10$^{-1}$ | -0.378 | 2.26 | 0.138 |
| | Control | IGE | 2.0(4.0) | 0.5(1.2) | -75.0 | 5.72 | **9.31x10$^{-8}$** | 5.70(1.67)x10$^{-1}$ | -0.565 | 3.42 | **0.005** |
| | TLE-HS | TLE- NEG | 1.0(2.0) | 1.0(2.0) | 0.0 | 0.12 | 1.000 | 4.76(14.00)x10$^{-2}$ | -0.028 | 0.34 | 0.997 |
| | TLE-HS | ETLE | 1.0(2.0) | 1.0(2.0) | 0.0 | 0.01 | 1.000 | -1.47(1.21)x10$^{-1}$ | 0.189 | -1.21 | 0.716 |
| | TLE-HS | IGE | 1.0(2.0) | 0.5(1.2) | -50.0 | 0.64 | 1.000 | 1.81(1.77)x10$^{-1}$ | 0.002 | 1.03 | 0.822 |
| | TLE- NEG | ETLE | 1.0(2.0) | 1.0(2.0) | 0.0 | 0.10 | 1.000 | -1.95(1.64)x10$^{-1}$ | 0.217 | -1.19 | 0.731 |
| | TLE- NEG | IGE | 1.0(2.0) | 0.5(1.2) | -50.0 | 0.42 | 1.000 | 1.34(2.08)x10$^{-1}$ | 0.030 | 0.64 | 0.963 |
| | ETLE | IGE | 1.0(2.0) | 0.5(1.2) | -50.0 | 0.51 | 1.000 | 3.28(1.95)x10$^{-1}$ | -0.187 | 1.68 | 0.411 |

Table S3.1: PVS Volume Fraction in the Basal Ganglia

| | Group | | median (IQR) (x10$^{-2}$) | | Univariate | | | General Linear Model | | | |
|---|---|---|---|---|---|---|---|---|---|---|---|
| | G1 | G2 | G1 | G2 | Δ% | Dunn | p-Holm | beta(sderr) | d(G) | z | p-Tukey |
| Basal Ganglia (BG) | Control | TLE-HS | 0.364(0.471) | 0.733(0.364) | 101.4 | 11.41 | **<1x10$^{-20}$** | 27.40(2.24)x10$^{-4}$ | 0.951 | 12.22 | **<1x10$^{-20}$** |
| | Control | TLE- NEG | 0.364(0.471) | 0.756(0.345) | 107.8 | 7.50 | **5.24x10$^{-13}$** | 30.00(3.69)x10$^{-4}$ | 1.039 | 8.11 | **3.55x10$^{-15}$** |
| | Control | ETLE | 0.364(0.471) | 0.872(0.315) | 139.6 | 9.02 | **1.70x10$^{-18}$** | 39.40(3.88)x10$^{-4}$ | 1.367 | 10.15 | **<1x10$^{-20}$** |
| | Control | IGE | 0.364(0.471) | 0.814(0.424) | 123.9 | 7.34 | **1.52x10$^{-12}$** | 31.90(3.86)x10$^{-4}$ | 1.108 | 8.27 | **1.33x10$^{-15}$** |
| | TLE-HS | TLE- NEG | 0.733(0.364) | 0.756(0.345) | 3.2 | 0.61 | 1.000 | 2.56(3.85)x10$^{-4}$ | 0.089 | 0.66 | 0.961 |
| | TLE-HS | ETLE | 0.733(0.364) | 0.872(0.315) | 19.0 | 2.31 | 0.124 | 12.00(4.13)x10$^{-4}$ | 0.416 | 2.90 | **0.028** |
| | TLE-HS | IGE | 0.733(0.364) | 0.814(0.424) | 11.2 | 0.75 | 1.000 | 4.53(4.10)x10$^{-4}$ | 0.157 | 1.11 | 0.790 |
| | TLE- NEG | ETLE | 0.756(0.345) | 0.872(0.315) | 15.3 | 1.39 | 0.819 | 9.44(5.07)x10$^{-4}$ | 0.327 | 1.86 | 0.319 |
| | TLE- NEG | IGE | 0.756(0.345) | 0.814(0.424) | 7.8 | 0.13 | 1.000 | 1.97(5.04)x10$^{-4}$ | 0.068 | 0.39 | 0.995 |
| | ETLE | IGE | 0.872(0.315) | 0.814(0.424) | -6.6 | 1.23 | 0.881 | -7.46(5.10)x10$^{-4}$ | -0.259 | -1.46 | 0.565 |
| BG excluding Thalami | Control | TLE-HS | 0.448(0.577) | 0.920(0.482) | 105.6 | 11.19 | **<1x10$^{-20}$** | 34.60(2.87)x10$^{-4}$ | 0.939 | 12.07 | **<1x10$^{-20}$** |
| | Control | TLE- NEG | 0.448(0.577) | 0.958(0.451) | 114.0 | 7.46 | **7.15x10$^{-13}$** | 38.40(4.72)x10$^{-4}$ | 1.042 | 8.13 | **3.11x10$^{-15}$** |
| | Control | ETLE | 0.448(0.577) | 1.121(0.400) | 150.4 | 9.25 | **2.01x10$^{-19}$** | 51.10(4.97)x10$^{-4}$ | 1.386 | 10.29 | **<1x10$^{-20}$** |
| | Control | IGE | 0.448(0.577) | 1.020(0.552) | 127.8 | 7.39 | **1.01x10$^{-12}$** | 40.50(4.94)x10$^{-4}$ | 1.098 | 8.19 | **1.67x10$^{-15}$** |
| | TLE-HS | TLE- NEG | 0.920(0.482) | 0.958(0.451) | 4.1 | 0.69 | 1.000 | 3.77(4.92)x10$^{-4}$ | 0.102 | 0.77 | 0.935 |
| | TLE-HS | ETLE | 0.920(0.482) | 1.121(0.400) | 21.8 | 2.65 | **0.048** | 16.50(5.29)x10$^{-4}$ | 0.447 | 3.12 | **0.014** |
| | TLE-HS | IGE | 0.920(0.482) | 1.020(0.552) | 10.8 | 0.92 | 1.000 | 5.85(5.24)x10$^{-4}$ | 0.159 | 1.12 | 0.784 |
| | TLE- NEG | ETLE | 0.958(0.451) | 1.121(0.400) | 17.0 | 1.60 | 0.547 | 12.70(6.49)x10$^{-4}$ | 0.345 | 1.96 | 0.269 |
| | TLE- NEG | IGE | 0.958(0.451) | 1.020(0.552) | 6.4 | 0.21 | 1.000 | 2.07(6.45)x10$^{-4}$ | 0.056 | 0.32 | 0.997 |
| | ETLE | IGE | 1.121(0.400) | 1.020(0.552) | -9.0 | 1.36 | 0.698 | -10.60(6.52)x10$^{-4}$ | -0.288 | -1.63 | 0.457 |
| Thalami | Control | TLE-HS | 0.017(0.064) | 0.049(0.096) | 184.4 | 4.49 | **7.00x10$^{-5}$** | -6.29(1.84)x10$^{+2}$ | 0.261 | -3.42 | **0.005** |
| | Control | TLE- NEG | 0.017(0.064) | 0.039(0.103) | 123.7 | 2.00 | 0.400 | -5.90(2.75)x10$^{+2}$ | 0.255 | -2.15 | 0.186 |
| | Control | ETLE | 0.017(0.064) | 0.019(0.070) | 12.5 | 0.50 | 1.000 | -2.49(3.87)x10$^{+2}$ | 0.079 | -0.64 | 0.965 |
| | Control | IGE | 0.017(0.064) | 0.024(0.092) | 41.2 | 1.78 | 0.525 | -6.61(3.27)x10$^{+2}$ | 0.236 | -2.02 | 0.238 |
| | TLE-HS | TLE- NEG | 0.049(0.096) | 0.039(0.103) | -21.4 | 0.67 | 1.000 | 3.91(26.70)x10$^{+1}$ | -0.006 | 0.15 | 1.000 |
| | TLE-HS | ETLE | 0.049(0.096) | 0.019(0.070) | -60.4 | 2.01 | 0.400 | 3.80(3.87)x10$^{+2}$ | -0.181 | 0.98 | 0.853 |
| | TLE-HS | IGE | 0.049(0.096) | 0.024(0.092) | -50.4 | 0.77 | 1.000 | -3.20(32.70)x10$^{+1}$ | -0.025 | -0.10 | 1.000 |
| | TLE- NEG | ETLE | 0.039(0.103) | 0.019(0.070) | -49.7 | 1.09 | 1.000 | 3.41(4.40)x10$^{+2}$ | -0.176 | 0.77 | 0.933 |
| | TLE- NEG | IGE | 0.039(0.103) | 0.024(0.092) | -36.9 | 0.10 | 1.000 | -7.11(38.70)x10$^{+1}$ | -0.019 | -0.18 | 1.000 |
| | ETLE | IGE | 0.019(0.070) | 0.024(0.092) | 25.4 | 0.97 | 1.000 | -4.12(4.62)x10$^{+2}$ | 0.157 | -0.89 | 0.892 |

Table S3.2: PVS total volume in the Basal Ganglia

| Region | Group | | median (IQR) | | Univariate | | | Generalised Linear Model | | | |
|---|---|---|---|---|---|---|---|---|---|---|---|
| | G1 | G2 | G1 | G2 | Δ% | Dunn | p-Holm | beta(sderr) | d(G) | z | p-Tukey |
| Basal Ganglia (BG) | Control | TLE-HS | 244(310) | 450(217) | 84.2 | 9.94 | **<1x$10^{-20}$** | 14.80(1.51)x$10^{+1}$ | 0.759 | 9.75 | **<1x$10^{-20}$** |
| | Control | TLE- NEG | 244(310) | 477(248) | 95.5 | 6.88 | **4.69x$10^{-11}$** | 17.20(2.49)x$10^{+1}$ | 0.885 | 6.91 | **1.87x$10^{-11}$** |
| | Control | ETLE | 244(310) | 538(280) | 120.5 | 8.60 | **7.15x$10^{-17}$** | 22.90(2.62)x$10^{+1}$ | 1.178 | 8.75 | **<1x$10^{-20}$** |
| | Control | IGE | 244(310) | 472(304) | 93.4 | 6.76 | **9.64x$10^{-11}$** | 19.00(2.60)x$10^{+1}$ | 0.975 | 7.28 | **1.17x$10^{-12}$** |
| | TLE-HS | TLE- NEG | 450(217) | 477(248) | 6.1 | 0.86 | 0.948 | 2.45(2.60)x$10^{+1}$ | 0.126 | 0.95 | 0.869 |
| | TLE-HS | ETLE | 450(217) | 538(280) | 19.7 | 2.72 | **0.039** | 8.15(2.79)x$10^{+1}$ | 0.419 | 2.92 | **0.026** |
| | TLE-HS | IGE | 450(217) | 472(304) | 5 | 1.00 | 0.948 | 4.20(2.76)x$10^{+1}$ | 0.216 | 1.52 | 0.527 |
| | TLE- NEG | ETLE | 477(248) | 538(280) | 12.8 | 1.52 | 0.637 | 5.70(3.42)x$10^{+1}$ | 0.293 | 1.67 | 0.434 |
| | TLE- NEG | IGE | 477(248) | 472(304) | -1 | 0.14 | 0.948 | 1.75(3.40)x$10^{+1}$ | 0.090 | 0.51 | 0.985 |
| | ETLE | IGE | 538(280) | 472(304) | -12.3 | 1.35 | 0.711 | -3.95(3.44)x$10^{+1}$ | -0.203 | -1.15 | 0.766 |
| BG excluding Thalami | Control | TLE-HS | 237(308) | 434(212) | 83.1 | 9.94 | **<1x$10^{-20}$** | 14.60(1.49)x$10^{+1}$ | 0.760 | 9.77 | **<1x$10^{-20}$** |
| | Control | TLE- NEG | 237(308) | 477(238) | 101.3 | 6.90 | **4.23x$10^{-11}$** | 17.00(2.46)x$10^{+1}$ | 0.887 | 6.92 | **1.83x$10^{-11}$** |
| | Control | ETLE | 237(308) | 529(274) | 123.2 | 8.69 | **3.34x$10^{-17}$** | 22.90(2.58)x$10^{+1}$ | 1.193 | 8.86 | **<1x$10^{-20}$** |
| | Control | IGE | 237(308) | 465(286) | 96.2 | 6.80 | **7.08x$10^{-11}$** | 18.80(2.57)x$10^{+1}$ | 0.980 | 7.32 | **1.62x$10^{-12}$** |
| | TLE-HS | TLE- NEG | 434(212) | 477(238) | 9.9 | 0.88 | 0.886 | 2.43(2.56)x$10^{+1}$ | 0.127 | 0.95 | 0.868 |
| | TLE-HS | ETLE | 434(212) | 529(274) | 21.9 | 2.81 | **0.030** | 8.31(2.75)x$10^{+1}$ | 0.433 | 3.02 | **0.019** |
| | TLE-HS | IGE | 434(212) | 465(286) | 7.1 | 1.05 | 0.886 | 4.22(2.72)x$10^{+1}$ | 0.220 | 1.55 | 0.509 |
| | TLE- NEG | ETLE | 477(238) | 529(274) | 10.9 | 1.58 | 0.570 | 5.88(3.37)x$10^{+1}$ | 0.307 | 1.74 | 0.386 |
| | TLE- NEG | IGE | 477(238) | 465(286) | -2.5 | 0.17 | 0.886 | 1.79(3.35)x$10^{+1}$ | 0.093 | 0.53 | 0.982 |
| | ETLE | IGE | 529(274) | 465(286) | -12.1 | 1.38 | 0.671 | -4.09(3.39)x$10^{+1}$ | -0.213 | -1.21 | 0.732 |
| Thalami | Control | TLE-HS | 2(9) | 6(12) | 182.8 | 3.84 | **0.001** | -3.65(1.33)x$10^{-2}$ | 0.211 | -2.75 | **0.043** |
| | Control | TLE- NEG | 2(9) | 4(13) | 88.6 | 1.72 | 0.771 | -3.90(1.97)x$10^{-2}$ | 0.239 | -1.98 | 0.260 |
| | Control | ETLE | 2(9) | 3(10) | 41.4 | 0.52 | 1.000 | -8.94(26.70)x$10^{-3}$ | 0.034 | -0.33 | 0.997 |
| | Control | IGE | 2(9) | 4(12) | 65 | 1.63 | 0.816 | -3.72(2.32)x$10^{-2}$ | 0.191 | -1.60 | 0.475 |
| | TLE-HS | TLE- NEG | 6(12) | 4(13) | -33.3 | 0.56 | 1.000 | -2.48(19.50)x$10^{-3}$ | 0.029 | -0.13 | 1.000 |
| | TLE-HS | ETLE | 6(12) | 3(10) | -50 | 1.62 | 0.816 | 2.76(2.70)x$10^{-2}$ | -0.177 | 1.02 | 0.834 |
| | TLE-HS | IGE | 6(12) | 4(12) | -41.7 | 0.55 | 1.000 | -6.60(235.00)x$10^{-4}$ | -0.020 | -0.03 | 1.000 |
| | TLE- NEG | ETLE | 4(13) | 3(10) | -25 | 0.87 | 1.000 | 3.01(3.08)x$10^{-2}$ | -0.205 | 0.98 | 0.856 |
| | TLE- NEG | IGE | 4(13) | 4(12) | -12.5 | 0.01 | 1.000 | 1.82(27.70)x$10^{-3}$ | -0.049 | 0.07 | 1.000 |
| | ETLE | IGE | 3(10) | 4(12) | 16.7 | 0.84 | 1.000 | -2.82(3.23)x$10^{-2}$ | 0.156 | -0.87 | 0.899 |

Table S3.3: PVS number in the Basal Ganglia

| | Group | | median (IQR) | | Univariate | | | Generalised Linear Model | | | |
|---|---|---|---|---|---|---|---|---|---|---|---|
| | G1 | G2 | G1 | G2 | Δ% | Dunn | p-Holm | beta(sderr) | d(G) | z | p-Tukey |
| Basal Ganglia (BG) | Control | TLE-HS | 25.0(19.0) | 36.0(11.0) | 44.0 | 10.38 | **<1x10$^{-20}$** | 69.30(8.07)x10$^{-1}$ | 0.668 | 8.58 | **<1x10$^{-20}$** |
| | Control | TLE- NEG | 25.0(19.0) | 36.0(15.0) | 44.0 | 6.73 | **1.55x10$^{-10}$** | 7.76(1.33)x10$^{+0}$ | 0.748 | 5.84 | **4.02x10$^{-8}$** |
| | Control | ETLE | 25.0(19.0) | 36.0(10.0) | 44.0 | 5.74 | **7.43x10$^{-8}$** | 10.60(1.40)x10$^{+0}$ | 1.023 | 7.60 | **1.42x10$^{-13}$** |
| | Control | IGE | 25.0(19.0) | 34.0(11.2) | 36.0 | 4.05 | **3.53x10$^{-4}$** | 8.14(1.39)x10$^{+0}$ | 0.785 | 5.86 | **6.55x10$^{-8}$** |
| | TLE-HS | TLE- NEG | 36.0(11.0) | 36.0(15.0) | 0.0 | 0.46 | 1.000 | 8.30(13.90)x10$^{-1}$ | 0.080 | 0.60 | 0.973 |
| | TLE-HS | ETLE | 36.0(11.0) | 36.0(10.0) | 0.0 | 0.25 | 1.000 | 3.69(1.49)x10$^{+0}$ | 0.356 | 2.48 | 0.087 |
| | TLE-HS | IGE | 36.0(11.0) | 34.0(11.2) | -5.6 | 1.83 | 0.399 | 1.21(1.47)x10$^{+0}$ | 0.117 | 0.82 | 0.918 |
| | TLE- NEG | ETLE | 36.0(15.0) | 36.0(10.0) | 0.0 | 0.56 | 1.000 | 2.86(1.83)x10$^{+0}$ | 0.276 | 1.57 | 0.498 |
| | TLE- NEG | IGE | 36.0(15.0) | 34.0(11.2) | -5.6 | 1.84 | 0.399 | 3.81(18.10)x10$^{-1}$ | 0.037 | 0.21 | 1.000 |
| | ETLE | IGE | 36.0(10.0) | 34.0(11.2) | -5.6 | 1.25 | 0.847 | -2.48(1.84)x10$^{+0}$ | -0.239 | -1.35 | 0.641 |
| BG excluding Thalami | Control | TLE-HS | 24.0(18.0) | 34.0(11.0) | 41.7 | 10.59 | **<1x10$^{-20}$** | 69.80(7.82)x10$^{-1}$ | 0.695 | 8.93 | **<1x10$^{-20}$** |
| | Control | TLE- NEG | 24.0(18.0) | 35.0(12.5) | 45.8 | 6.98 | **2.58x10$^{-11}$** | 7.99(1.29)x10$^{+0}$ | 0.795 | 6.21 | **2.64x10$^{-9}$** |
| | Control | ETLE | 24.0(18.0) | 35.0(10.0) | 45.8 | 6.06 | **1.07x10$^{-8}$** | 10.80(1.35)x10$^{+0}$ | 1.072 | 7.96 | **1.10x10$^{-14}$** |
| | Control | IGE | 24.0(18.0) | 33.0(10.2) | 37.5 | 4.10 | **2.92x10$^{-4}$** | 7.85(1.35)x10$^{+0}$ | 0.782 | 5.83 | **2.32x10$^{-8}$** |
| | TLE-HS | TLE- NEG | 34.0(11.0) | 35.0(12.5) | 2.9 | 0.58 | 1.000 | 1.00(1.34)x10$^{+0}$ | 0.100 | 0.75 | 0.940 |
| | TLE-HS | ETLE | 34.0(11.0) | 35.0(10.0) | 2.9 | 0.06 | 1.000 | 3.79(1.44)x10$^{+0}$ | 0.377 | 2.63 | 0.059 |
| | TLE-HS | IGE | 34.0(11.0) | 33.0(10.2) | -2.9 | 1.91 | 0.284 | 8.68(14.30)x10$^{-1}$ | 0.086 | 0.61 | 0.971 |
| | TLE- NEG | ETLE | 35.0(12.5) | 35.0(10.0) | 0.0 | 0.51 | 1.000 | 2.79(1.77)x10$^{+0}$ | 0.277 | 1.58 | 0.492 |
| | TLE- | IGE | 35.0(12.5) | 33.0(10.2) | -5.7 | 1.99 | 0.279 | -1.37(17.60)x10$^{-1}$ | -0.014 | -0.08 | 1.000 |
| | ETLE | IGE | 35.0(10.0) | 33.0(10.2) | -5.7 | 1.45 | 0.583 | -2.92(1.78)x10$^{+0}$ | -0.291 | -1.64 | 0.447 |
| Thalami | Control | TLE-HS | 1.0(2.0) | 1.0(3.0) | 0.0 | 2.82 | **0.048** | -8.15(5.15)x10$^{-2}$ | 0.111 | -1.58 | 0.488 |
| | Control | TLE- NEG | 1.0(2.0) | 1.0(3.0) | 0.0 | 0.77 | 1.000 | -3.36(8.64)x10$^{-2}$ | 0.040 | -0.39 | 0.995 |
| | Control | ETLE | 1.0(2.0) | 1.0(2.0) | 0.0 | 0.11 | 1.000 | -2.01(10.40)x10$^{-2}$ | 0.025 | -0.19 | 1.000 |
| | Control | IGE | 1.0(2.0) | 2.0(2.2) | 100.0 | 1.90 | 0.512 | -19.90(8.25)x10$^{-2}$ | 0.300 | -2.41 | 0.102 |
| | TLE-HS | TLE- NEG | 1.0(3.0) | 1.0(3.0) | 0.0 | 0.88 | 1.000 | 4.79(8.79)x10$^{-2}$ | -0.072 | 0.54 | 0.981 |
| | TLE-HS | ETLE | 1.0(3.0) | 1.0(2.0) | 0.0 | 1.46 | 1.000 | 6.14(10.60)x10$^{-2}$ | -0.086 | 0.58 | 0.976 |
| | TLE-HS | IGE | 1.0(3.0) | 2.0(2.2) | 100.0 | 0.27 | 1.000 | -11.80(8.55)x10$^{-2}$ | 0.189 | -1.38 | 0.623 |
| | TLE- NEG | ETLE | 1.0(3.0) | 1.0(2.0) | 0.0 | 0.49 | 1.000 | 1.35(12.70)x10$^{-2}$ | -0.014 | 0.11 | 1.000 |
| | TLE- NEG | IGE | 1.0(3.0) | 2.0(2.2) | 100.0 | 0.90 | 1.000 | -1.66(1.10)x10$^{-1}$ | 0.261 | -1.50 | 0.542 |
| | ETLE | IGE | 1.0(2.0) | 2.0(2.2) | 100.0 | 1.36 | 1.000 | -1.79(1.22)x10$^{-1}$ | 0.275 | -1.47 | 0.560 |

Table S4.0.1: Median and mean asymmetry measures for each group. Measures where the mean is significantly different from zero are highlighted in bold.

| | | PVS-Volume Fraction | | | PVS-Volume | | | PVS-Number | | |
|---|---|---|---|---|---|---|---|---|---|---|
| | group | median(iqr) (x10$^{-2}$) | mean(ci) (x10$^{-2}$) | p(t) | median(iqr) (x10$^{-2}$) | mean(ci) (x10$^{-2}$) | p(t) | median(iqr) (x10$^{-2}$) | mean(ci) (x10$^{-2}$) | p(t) |
| WM | Control | -0.18(16.27) | 0.47(1.40) | 0.508 | -0.36(16.43) | 0.42(1.40) | 0.561 | -1.23(12.96) | -0.72(1.18) | 0.235 |
| | TLE-HS | -1.28(14.16) | -2.41(1.70) | **0.006** | -2.27(14.27) | -3.60(1.76) | **$<10^{-3}$** | -2.07(13.70) | -2.51(1.39) | **$<10^{-3}$** |
| | TLE-NEG | 0.00(7.15) | 0.75(1.96) | 0.447 | 0.00(7.81) | 0.73(1.98) | 0.463 | 0.00(4.35) | 1.18(1.68) | 0.165 |
| | ETLE | -1.90(19.98) | -2.70(3.51) | 0.129 | -4.77(20.41) | -3.16(3.56) | 0.081 | 0.72(15.98) | -1.53(2.68) | 0.259 |
| | IGE | 0.79(13.99) | 1.38(2.98) | 0.358 | 0.57(14.19) | 1.27(2.97) | 0.397 | 1.76(11.96) | 0.03(2.39) | 0.982 |
| Frontal | Control | 0.55(23.80) | 1.94(2.18) | 0.081 | 1.26(23.23) | 2.74(2.18) | **0.014** | 0.00(20.86) | 1.21(1.86) | 0.202 |
| | TLE-HS | -0.92(19.46) | -3.68(2.33) | **0.002** | -1.50(19.79) | -4.19(2.35) | **0.001** | -1.96(14.84) | -2.84(1.83) | **0.003** |
| | TLE-NEG | 0.00(7.98) | 1.13(3.46) | 0.518 | 0.00(6.92) | 1.19(3.48) | 0.497 | 0.00(6.69) | -0.33(2.59) | 0.800 |
| | ETLE | 4.26(26.45) | 0.15(6.42) | 0.963 | 3.79(26.38) | 0.03(6.45) | 0.992 | 0.00(20.60) | -1.90(4.72) | 0.424 |
| | IGE | 4.88(23.17) | 5.44(4.68) | **0.024** | 5.77(25.80) | 6.36(4.69) | **0.009** | 2.68(18.66) | 2.76(3.57) | 0.128 |
| Parietal | Control | -4.26(26.91) | -2.18(2.25) | 0.057 | -3.06(26.41) | -1.10(2.24) | 0.334 | -3.23(21.43) | -1.60(1.87) | 0.094 |
| | TLE-HS | -2.35(22.99) | -3.09(2.88) | **0.035** | -3.03(22.02) | -3.92(2.88) | **0.008** | 0.00(22.74) | -2.13(2.34) | 0.074 |
| | TLE-NEG | 0.00(14.99) | 2.26(3.70) | 0.227 | 0.00(13.68) | 2.16(3.56) | 0.230 | 0.00(7.76) | 3.27(2.73) | **0.019** |
| | ETLE | -3.45(26.65) | -3.70(5.05) | 0.148 | -4.45(24.52) | -4.51(5.02) | 0.078 | -1.59(15.65) | -2.44(4.56) | 0.288 |
| | IGE | -4.37(33.88) | -1.24(7.50) | 0.742 | -2.36(32.24) | -0.35(7.50) | 0.926 | 0.00(21.98) | -2.52(5.98) | 0.403 |
| Temporal | Control | 0.00(76.32) | -6.81(5.26) | **0.011** | 0.00(75.00) | -8.14(5.25) | **0.002** | 0.00(44.44) | -7.50(4.68) | **0.002** |
| | TLE-HS | -2.18(80.61) | -9.29(7.10) | **0.011** | -4.17(82.02) | -10.76(7.11) | **0.003** | 0.00(55.09) | -7.43(6.41) | **0.023** |
| | TLE-NEG | 0.00(8.88) | -1.14(11.41) | 0.843 | 0.00(10.56) | -1.31(11.43) | 0.820 | 0.00(11.33) | 2.07(10.69) | 0.700 |
| | ETLE | 0.00(65.49) | 2.25(14.61) | 0.759 | 0.00(66.67) | 2.25(14.61) | 0.759 | 0.00(53.33) | 1.62(13.57) | 0.812 |
| | IGE | 0.00(84.05) | -7.39(16.06) | 0.361 | 0.00(86.05) | -8.16(16.05) | 0.314 | 0.00(46.67) | -4.72(14.83) | 0.527 |
| Occipital | Control | 0.00(93.31) | 5.84(5.90) | 0.052 | 0.00(93.33) | 6.17(5.91) | **0.041** | 0.00(66.67) | 5.90(5.45) | **0.034** |
| | TLE-HS | 0.00(12.96) | 0.85(7.41) | 0.821 | 0.00(10.22) | 0.86(7.41) | 0.820 | 0.00(0.00) | 1.11(7.16) | 0.761 |
| | TLE-NEG | 0.00(0.00) | 5.85(11.87) | 0.329 | 0.00(0.00) | 5.93(11.82) | 0.321 | 0.00(0.00) | 2.97(11.61) | 0.612 |
| | ETLE | 0.00(61.53) | -14.04(15.21) | **0.070** | 0.00(64.71) | -13.89(15.22) | 0.073 | 0.00(50.00) | -12.11(15.08) | 0.114 |
| | IGE | 0.00(14.78) | 1.09(16.00) | 0.892 | 0.00(11.58) | 1.33(15.97) | 0.868 | 0.00(0.00) | 1.56(15.39) | 0.840 |

Table S4.1: PVS Volume Fraction asymmetry in the white matter (WM)

| | Group | | median (IQR) | | Univariate | | Generalised Linear Model | | | |
|---|---|---|---|---|---|---|---|---|---|---|
| | G1 | G2 | G1 | G2 | Dunn | p-Holm | beta(sderr) | d(G) | z | p-Tukey |
| WM | Control | TLE-HS | -0.002(0.163) | -0.013(0.142) | 2.18 | 0.296 | -2.61(1.12)x10$^{-2}$ | -0.182 | -2.33 | 0.124 |
| | Control | TLE- NEG | -0.002(0.163) | 0.000(0.072) | 0.64 | 1.000 | 5.86(18.40)x10$^{-3}$ | 0.041 | 0.32 | 0.998 |
| | Control | ETLE | -0.002(0.163) | -0.019(0.200) | 1.43 | 0.770 | -3.25(1.93)x10$^{-2}$ | -0.226 | -1.68 | 0.426 |
| | Control | IGE | -0.002(0.163) | 0.008(0.140) | 0.86 | 1.000 | 6.17(19.20)x10$^{-3}$ | 0.043 | 0.32 | 0.997 |
| | TLE-HS | TLE- NEG | -0.013(0.142) | 0.000(0.072) | 1.86 | 0.506 | 3.19(1.92)x10$^{-2}$ | 0.222 | 1.67 | 0.435 |
| | TLE-HS | ETLE | -0.013(0.142) | -0.019(0.200) | 0.16 | 1.000 | -6.41(20.60)x10$^{-3}$ | -0.045 | -0.31 | 0.998 |
| | TLE-HS | IGE | -0.013(0.142) | 0.008(0.140) | 2.02 | 0.391 | 3.22(2.04)x10$^{-2}$ | 0.224 | 1.58 | 0.489 |
| | TLE- NEG | ETLE | 0.000(0.072) | -0.019(0.200) | 1.57 | 0.694 | -3.83(2.53)x10$^{-2}$ | -0.267 | -1.52 | 0.530 |
| | TLE- NEG | IGE | 0.000(0.072) | 0.008(0.140) | 0.19 | 1.000 | 3.03(251.00)x10$^{-4}$ | 0.002 | 0.01 | 1.000 |
| | ETLE | IGE | -0.019(0.200) | 0.008(0.140) | 1.72 | 0.597 | 3.86(2.54)x10$^{-2}$ | 0.269 | 1.52 | 0.527 |
| WM - Frontal | Control | TLE-HS | 0.005(0.238) | -0.009(0.195) | 2.53 | 0.103 | -5.19(1.71)x10$^{-2}$ | -0.236 | -3.03 | **0.019** |
| | Control | TLE- NEG | 0.005(0.238) | 0.000(0.080) | 0.25 | 1.000 | -3.33(28.20)x10$^{-3}$ | -0.015 | -0.12 | 1.000 |
| | Control | ETLE | 0.005(0.238) | 0.043(0.265) | 0.07 | 1.000 | -2.13(2.96)x10$^{-2}$ | -0.097 | -0.72 | 0.948 |
| | Control | IGE | 0.005(0.238) | 0.049(0.232) | 1.69 | 0.729 | 3.04(2.95)x10$^{-2}$ | 0.138 | 1.03 | 0.828 |
| | TLE-HS | TLE- NEG | -0.009(0.195) | 0.000(0.080) | 1.69 | 0.729 | 4.85(2.94)x10$^{-2}$ | 0.221 | 1.65 | 0.443 |
| | TLE-HS | ETLE | -0.009(0.195) | 0.043(0.265) | 1.33 | 1.000 | 3.06(3.15)x10$^{-2}$ | 0.139 | 0.97 | 0.859 |
| | TLE-HS | IGE | -0.009(0.195) | 0.049(0.232) | 3.01 | **0.027** | 8.23(3.13)x10$^{-2}$ | 0.374 | 2.63 | 0.059 |
| | TLE- NEG | ETLE | 0.000(0.080) | 0.043(0.265) | 0.24 | 1.000 | -1.80(3.87)x10$^{-2}$ | -0.082 | -0.46 | 0.990 |
| | TLE- NEG | IGE | 0.000(0.080) | 0.049(0.232) | 1.12 | 1.000 | 3.38(3.85)x10$^{-2}$ | 0.153 | 0.88 | 0.897 |
| | ETLE | IGE | 0.043(0.265) | 0.049(0.232) | 1.33 | 1.000 | 5.17(3.89)x10$^{-2}$ | 0.235 | 1.33 | 0.654 |
| WM - Parietal | Control | TLE-HS | -0.043(0.269) | -0.024(0.230) | 0.62 | 1.000 | -1.08(1.88)x10$^{-2}$ | -0.045 | -0.57 | 0.977 |
| | Control | TLE- NEG | -0.043(0.269) | 0.000(0.150) | 2.27 | 0.229 | 4.27(3.09)x10$^{-2}$ | 0.177 | 1.38 | 0.622 |
| | Control | ETLE | -0.043(0.269) | -0.034(0.266) | 0.28 | 1.000 | -1.24(3.25)x10$^{-2}$ | -0.051 | -0.38 | 0.995 |
| | Control | IGE | -0.043(0.269) | -0.044(0.339) | 0.29 | 1.000 | 1.11(3.24)x10$^{-2}$ | 0.046 | 0.34 | 0.997 |
| | TLE-HS | TLE- NEG | -0.024(0.230) | 0.000(0.150) | 1.81 | 0.562 | 5.35(3.22)x10$^{-2}$ | 0.221 | 1.66 | 0.439 |
| | TLE-HS | ETLE | -0.024(0.230) | -0.034(0.266) | 0.61 | 1.000 | -1.61(34.60)x10$^{-3}$ | -0.007 | -0.05 | 1.000 |
| | TLE-HS | IGE | -0.024(0.230) | -0.044(0.339) | 0.07 | 1.000 | 2.19(3.43)x10$^{-2}$ | 0.091 | 0.64 | 0.966 |
| | TLE- NEG | ETLE | 0.000(0.150) | -0.034(0.266) | 1.90 | 0.516 | -5.51(4.25)x10$^{-2}$ | -0.228 | -1.30 | 0.676 |
| | TLE- NEG | IGE | 0.000(0.150) | -0.044(0.339) | 1.46 | 1.000 | -3.15(4.22)x10$^{-2}$ | -0.131 | -0.75 | 0.941 |
| | ETLE | IGE | -0.034(0.266) | -0.044(0.339) | 0.43 | 1.000 | 2.35(4.27)x10$^{-2}$ | 0.097 | 0.55 | 0.980 |
| WM - Temporal | Control | TLE-HS | 0.000(0.763) | -0.022(0.806) | 0.66 | 1.000 | -2.20(4.54)x10$^{-2}$ | -0.038 | -0.49 | 0.988 |
| | Control | TLE- NEG | 0.000(0.763) | 0.000(0.089) | 0.44 | 1.000 | 5.98(7.47)x10$^{-2}$ | 0.103 | 0.80 | 0.924 |
| | Control | ETLE | 0.000(0.763) | 0.000(0.655) | 1.08 | 1.000 | 8.94(7.85)x10$^{-2}$ | 0.153 | 1.14 | 0.772 |
| | Control | IGE | 0.000(0.763) | 0.000(0.840) | 0.22 | 1.000 | -8.78(78.10)x10$^{-3}$ | -0.015 | -0.11 | 1.000 |
| | TLE-HS | TLE- NEG | -0.022(0.806) | 0.000(0.089) | 0.80 | 1.000 | 8.18(7.78)x10$^{-2}$ | 0.140 | 1.05 | 0.819 |
| | TLE-HS | ETLE | -0.022(0.806) | 0.000(0.655) | 1.39 | 1.000 | 11.10(8.36)x10$^{-2}$ | 0.191 | 1.33 | 0.652 |
| | TLE-HS | IGE | -0.022(0.806) | 0.000(0.840) | 0.16 | 1.000 | 1.32(8.28)x10$^{-2}$ | 0.023 | 0.16 | 1.000 |
| | TLE- NEG | ETLE | 0.000(0.089) | 0.000(0.655) | 0.50 | 1.000 | 2.95(10.30)x10$^{-2}$ | 0.051 | 0.29 | 0.998 |
| | TLE- NEG | IGE | 0.000(0.089) | 0.000(0.840) | 0.49 | 1.000 | -6.86(10.20)x10$^{-2}$ | -0.118 | -0.67 | 0.959 |
| | ETLE | IGE | 0.000(0.655) | 0.000(0.840) | 0.97 | 1.000 | -9.81(10.30)x10$^{-2}$ | -0.168 | -0.95 | 0.867 |
| WM - Occipital | Control | TLE-HS | 0.000(0.933) | 0.000(0.130) | 1.27 | 1.000 | -5.34(4.89)x10$^{-2}$ | -0.085 | -1.09 | 0.798 |
| | Control | TLE- NEG | 0.000(0.933) | 0.000(0.000) | 0.15 | 1.000 | -3.36(80.60)x10$^{-3}$ | -0.005 | -0.04 | 1.000 |
| | Control | ETLE | 0.000(0.933) | 0.000(0.615) | 2.29 | 0.223 | -18.50(8.47)x10$^{-2}$ | -0.294 | -2.19 | 0.172 |
| | Control | IGE | 0.000(0.933) | 0.000(0.148) | 0.65 | 1.000 | -4.41(8.43)x10$^{-2}$ | -0.070 | -0.52 | 0.984 |
| | TLE-HS | TLE- NEG | 0.000(0.130) | 0.000(0.000) | 0.58 | 1.000 | 5.00(8.40)x10$^{-2}$ | 0.080 | 0.60 | 0.973 |
| | TLE-HS | ETLE | 0.000(0.130) | 0.000(0.615) | 1.48 | 1.000 | -13.20(9.02)x10$^{-2}$ | -0.210 | -1.46 | 0.567 |
| | TLE-HS | IGE | 0.000(0.130) | 0.000(0.148) | 0.07 | 1.000 | 9.31(89.40)x10$^{-3}$ | 0.015 | 0.10 | 1.000 |
| | TLE- NEG | ETLE | 0.000(0.000) | 0.000(0.615) | 1.65 | 0.895 | -1.82(1.11)x10$^{-1}$ | -0.289 | -1.64 | 0.448 |
| | TLE- NEG | IGE | 0.000(0.000) | 0.000(0.148) | 0.39 | 1.000 | -4.07(11.00)x10$^{-2}$ | -0.065 | -0.37 | 0.996 |
| | ETLE | IGE | 0.000(0.615) | 0.000(0.148) | 1.22 | 1.000 | 1.41(1.11)x10$^{-1}$ | 0.224 | 1.27 | 0.693 |

Table S4.2: PVS-Volume asymmetry in the white matter (WM)

| | Group | | median (IQR) | | Univariate | | Generalised Linear Model | | | |
|---|---|---|---|---|---|---|---|---|---|---|
| | G1 | G2 | G1 | G2 | Dunn | p-Holm | beta(sderr) | d(G) | z | p-Tukey |
| WM | Control | TLE-HS | -0.004(0.164) | -0.023(0.143) | 3.18 | **0.015** | -3.72(1.13)x10$^{-2}$ | -0.256 | -3.29 | **0.008** |
| | Control | TLE- NEG | -0.004(0.164) | 0.000(0.078) | 0.73 | 1.000 | 6.44(18.60)x10$^{-3}$ | 0.044 | 0.35 | 0.997 |
| | Control | ETLE | -0.004(0.164) | -0.048(0.204) | 1.70 | 0.450 | -3.68(1.95)x10$^{-2}$ | -0.254 | -1.88 | 0.308 |
| | Control | IGE | -0.004(0.164) | 0.006(0.142) | 0.82 | 1.000 | 5.41(19.40)x10$^{-3}$ | 0.037 | 0.28 | 0.999 |
| | TLE-HS | TLE- NEG | -0.023(0.143) | 0.000(0.078) | 2.53 | 0.101 | 4.36(1.94)x10$^{-2}$ | 0.301 | 2.25 | 0.149 |
| | TLE-HS | ETLE | -0.023(0.143) | -0.048(0.204) | 0.14 | 1.000 | 3.73(208.00)x10$^{-4}$ | 0.003 | 0.02 | 1.000 |
| | TLE-HS | IGE | -0.023(0.143) | 0.006(0.142) | 2.54 | 0.101 | 4.26(2.06)x10$^{-2}$ | 0.293 | 2.07 | 0.219 |
| | TLE- NEG | ETLE | 0.000(0.078) | -0.048(0.204) | 1.85 | 0.407 | -4.32(2.55)x10$^{-2}$ | -0.298 | -1.69 | 0.417 |
| | TLE- NEG | IGE | 0.000(0.078) | 0.006(0.142) | 0.09 | 1.000 | -1.03(25.40)x10$^{-3}$ | -0.007 | -0.04 | 1.000 |
| | ETLE | IGE | -0.048(0.204) | 0.006(0.142) | 1.89 | 0.407 | 4.22(2.57)x10$^{-2}$ | 0.291 | 1.64 | 0.448 |
| WM - Frontal | Control | TLE-HS | 0.013(0.232) | -0.015(0.198) | 3.38 | **0.007** | -6.53(1.72)x10$^{-2}$ | -0.296 | -3.80 | **0.001** |
| | Control | TLE- NEG | 0.013(0.232) | 0.000(0.069) | 0.15 | 1.000 | -1.10(2.83)x10$^{-2}$ | -0.050 | -0.39 | 0.995 |
| | Control | ETLE | 0.013(0.232) | 0.038(0.264) | 0.43 | 1.000 | -3.00(2.97)x10$^{-2}$ | -0.136 | -1.01 | 0.841 |
| | Control | IGE | 0.013(0.232) | 0.058(0.258) | 1.76 | 0.590 | 3.20(2.96)x10$^{-2}$ | 0.145 | 1.08 | 0.802 |
| | TLE-HS | TLE- NEG | -0.015(0.198) | 0.000(0.069) | 1.79 | 0.590 | 5.43(2.95)x10$^{-2}$ | 0.246 | 1.84 | 0.331 |
| | TLE-HS | ETLE | -0.015(0.198) | 0.038(0.264) | 1.46 | 0.707 | 3.53(3.17)x10$^{-2}$ | 0.160 | 1.12 | 0.784 |
| | TLE-HS | IGE | -0.015(0.198) | 0.058(0.258) | 3.54 | **0.004** | 9.73(3.14)x10$^{-2}$ | 0.441 | 3.10 | **0.015** |
| | TLE- NEG | ETLE | 0.000(0.069) | 0.038(0.264) | 0.21 | 1.000 | -1.89(3.88)x10$^{-2}$ | -0.086 | -0.49 | 0.987 |
| | TLE- NEG | IGE | 0.000(0.069) | 0.058(0.258) | 1.47 | 0.707 | 4.30(3.86)x10$^{-2}$ | 0.195 | 1.11 | 0.785 |
| | ETLE | IGE | 0.038(0.264) | 0.058(0.258) | 1.65 | 0.597 | 6.20(3.91)x10$^{-2}$ | 0.281 | 1.59 | 0.485 |
| WM - Parietal | Control | TLE-HS | -0.031(0.264) | -0.030(0.220) | 0.57 | 1.000 | -3.01(1.87)x10$^{-2}$ | -0.125 | -1.61 | 0.472 |
| | Control | TLE- NEG | -0.031(0.264) | 0.000(0.137) | 1.73 | 0.662 | 3.06(3.08)x10$^{-2}$ | 0.127 | 0.99 | 0.848 |
| | Control | ETLE | -0.031(0.264) | -0.045(0.245) | 1.06 | 1.000 | -3.05(3.24)x10$^{-2}$ | -0.127 | -0.94 | 0.871 |
| | Control | IGE | -0.031(0.264) | -0.024(0.322) | 0.22 | 1.000 | 9.52(32.20)x10$^{-3}$ | 0.040 | 0.30 | 0.998 |
| | TLE-HS | TLE- NEG | -0.030(0.220) | 0.000(0.137) | 1.98 | 0.429 | 6.07(3.21)x10$^{-2}$ | 0.252 | 1.89 | 0.305 |
| | TLE-HS | ETLE | -0.030(0.220) | -0.045(0.245) | 0.70 | 1.000 | -4.75(345.00)x10$^{-4}$ | -0.002 | -0.01 | 1.000 |
| | TLE-HS | IGE | -0.030(0.220) | -0.024(0.322) | 0.53 | 1.000 | 3.96(3.42)x10$^{-2}$ | 0.165 | 1.16 | 0.760 |
| | TLE- NEG | ETLE | 0.000(0.137) | -0.045(0.245) | 2.11 | 0.351 | -6.11(4.23)x10$^{-2}$ | -0.254 | -1.44 | 0.578 |
| | TLE- NEG | IGE | 0.000(0.137) | -0.024(0.322) | 1.11 | 1.000 | -2.11(4.21)x10$^{-2}$ | -0.088 | -0.50 | 0.986 |
| | ETLE | IGE | -0.045(0.245) | -0.024(0.322) | 0.97 | 1.000 | 4.01(4.25)x10$^{-2}$ | 0.167 | 0.94 | 0.871 |
| WM - Temporal | Control | TLE-HS | 0.000(0.750) | -0.042(0.820) | 0.68 | 1.000 | -2.22(4.53)x10$^{-2}$ | -0.038 | -0.49 | 0.987 |
| | Control | TLE- NEG | 0.000(0.750) | 0.000(0.106) | 0.72 | 1.000 | 7.26(7.46)x10$^{-2}$ | 0.125 | 0.97 | 0.857 |
| | Control | ETLE | 0.000(0.750) | 0.000(0.667) | 1.36 | 1.000 | 10.10(7.85)x10$^{-2}$ | 0.173 | 1.29 | 0.682 |
| | Control | IGE | 0.000(0.750) | 0.000(0.861) | 0.16 | 1.000 | -4.36(78.00)x10$^{-3}$ | -0.007 | -0.06 | 1.000 |
| | TLE-HS | TLE- NEG | -0.042(0.820) | 0.000(0.106) | 1.08 | 1.000 | 9.49(7.78)x10$^{-2}$ | 0.163 | 1.22 | 0.723 |
| | TLE-HS | ETLE | -0.042(0.820) | 0.000(0.667) | 1.67 | 0.948 | 12.30(8.35)x10$^{-2}$ | 0.211 | 1.47 | 0.558 |
| | TLE-HS | IGE | -0.042(0.820) | 0.000(0.861) | 0.22 | 1.000 | 1.79(8.28)x10$^{-2}$ | 0.031 | 0.22 | 0.999 |
| | TLE- NEG | ETLE | 0.000(0.106) | 0.000(0.667) | 0.51 | 1.000 | 2.83(10.20)x10$^{-2}$ | 0.049 | 0.28 | 0.999 |
| | TLE- NEG | IGE | 0.000(0.106) | 0.000(0.861) | 0.66 | 1.000 | -7.70(10.20)x10$^{-2}$ | -0.132 | -0.76 | 0.938 |
| | ETLE | IGE | 0.000(0.667) | 0.000(0.861) | 1.14 | 1.000 | -1.05(1.03)x10$^{-1}$ | -0.181 | -1.02 | 0.834 |
| WM - Occipital | Control | TLE-HS | 0.000(0.933) | 0.000(0.102) | 1.25 | 1.000 | -5.65(4.90)x10$^{-2}$ | -0.090 | -1.15 | 0.762 |
| | Control | TLE- NEG | 0.000(0.933) | 0.000(0.000) | 0.02 | 1.000 | -5.78(80.60)x10$^{-3}$ | -0.009 | -0.07 | 1.000 |
| | Control | ETLE | 0.000(0.933) | 0.000(0.647) | 2.34 | 0.191 | -18.70(8.48)x10$^{-2}$ | -0.297 | -2.21 | 0.164 |
| | Control | IGE | 0.000(0.933) | 0.000(0.116) | 0.70 | 1.000 | -4.49(8.43)x10$^{-2}$ | -0.071 | -0.53 | 0.982 |
| | TLE-HS | TLE- NEG | 0.000(0.102) | 0.000(0.000) | 0.70 | 1.000 | 5.08(8.40)x10$^{-2}$ | 0.081 | 0.60 | 0.972 |
| | TLE-HS | ETLE | 0.000(0.102) | 0.000(0.647) | 1.55 | 0.974 | -13.10(9.02)x10$^{-2}$ | -0.208 | -1.45 | 0.576 |
| | TLE-HS | IGE | 0.000(0.102) | 0.000(0.116) | 0.02 | 1.000 | 1.16(8.94)x10$^{-2}$ | 0.018 | 0.13 | 1.000 |
| | TLE- NEG | ETLE | 0.000(0.000) | 0.000(0.647) | 1.79 | 0.662 | -1.81(1.11)x10$^{-1}$ | -0.288 | -1.64 | 0.451 |
| | TLE- NEG | IGE | 0.000(0.000) | 0.000(0.116) | 0.53 | 1.000 | -3.91(11.00)x10$^{-2}$ | -0.062 | -0.36 | 0.996 |
| | ETLE | IGE | 0.000(0.647) | 0.000(0.116) | 1.23 | 1.000 | 1.42(1.11)x10$^{-1}$ | 0.226 | 1.28 | 0.687 |

Table S4.3: PVS number asymmetry in the white matter (WM)

| | Group | | median (IQR) | | Univariate | | General Linear Model | | | |
|---|---|---|---|---|---|---|---|---|---|---|
| | G1 | G2 | G1 | G2 | Dunn | p-Holm | beta(sderr) | d(G) | z | p-Tukey |
| WM | Control | TLE-HS | -0.012(0.130) | -0.021(0.137) | 1.53 | 0.881 | $-18.30(9.28)\times10^{-3}$ | -0.154 | -1.98 | 0.260 |
| | Control | TLE- NEG | -0.012(0.130) | 0.000(0.043) | 1.74 | 0.648 | $1.87(1.53)\times10^{-2}$ | 0.157 | 1.22 | 0.722 |
| | Control | EXTRA | -0.012(0.130) | 0.007(0.160) | 0.05 | 1.000 | $-5.34(16.10)\times10^{-3}$ | -0.045 | -0.33 | 0.997 |
| | Control | IGE | -0.012(0.130) | 0.018(0.120) | 1.11 | 1.000 | $7.75(16.00)\times10^{-3}$ | 0.065 | 0.49 | 0.988 |
| | TLE-HS | TLE- NEG | -0.021(0.137) | 0.000(0.043) | 2.54 | 0.111 | $3.70(1.59)\times10^{-2}$ | 0.310 | 2.32 | 0.127 |
| | TLE-HS | EXTRA | -0.021(0.137) | 0.007(0.160) | 0.80 | 1.000 | $1.30(1.71)\times10^{-2}$ | 0.109 | 0.76 | 0.937 |
| | TLE-HS | IGE | -0.021(0.137) | 0.018(0.120) | 1.90 | 0.516 | $2.61(1.69)\times10^{-2}$ | 0.219 | 1.54 | 0.516 |
| | TLE- NEG | EXTRA | 0.000(0.043) | 0.007(0.160) | 1.34 | 1.000 | $-2.40(2.10)\times10^{-2}$ | -0.201 | -1.15 | 0.768 |
| | TLE- NEG | IGE | 0.000(0.043) | 0.018(0.120) | 0.43 | 1.000 | $-1.09(2.08)\times10^{-2}$ | -0.092 | -0.52 | 0.983 |
| | EXTRA | IGE | 0.007(0.160) | 0.018(0.120) | 0.88 | 1.000 | $1.31(2.11)\times10^{-2}$ | 0.110 | 0.62 | 0.969 |
| WM - Frontal | Control | TLE-HS | 0.000(0.209) | -0.020(0.148) | 2.53 | 0.103 | $-3.95(1.41)\times10^{-2}$ | -0.219 | -2.81 | **0.036** |
| | Control | TLE- NEG | 0.000(0.209) | 0.000(0.067) | 0.05 | 1.000 | $-1.42(2.32)\times10^{-2}$ | -0.079 | -0.61 | 0.971 |
| | Control | EXTRA | 0.000(0.209) | 0.000(0.206) | 0.77 | 1.000 | $-2.86(2.44)\times10^{-2}$ | -0.158 | -1.17 | 0.752 |
| | Control | IGE | 0.000(0.209) | 0.027(0.187) | 1.24 | 1.000 | $1.45(2.42)\times10^{-2}$ | 0.080 | 0.60 | 0.973 |
| | TLE-HS | TLE- NEG | -0.020(0.148) | 0.000(0.067) | 1.40 | 1.000 | $2.53(2.41)\times10^{-2}$ | 0.140 | 1.05 | 0.820 |
| | TLE-HS | EXTRA | -0.020(0.148) | 0.000(0.206) | 0.66 | 1.000 | $1.10(2.59)\times10^{-2}$ | 0.061 | 0.42 | 0.993 |
| | TLE-HS | IGE | -0.020(0.148) | 0.027(0.187) | 2.58 | 0.098 | $5.40(2.57)\times10^{-2}$ | 0.299 | 2.10 | 0.204 |
| | TLE- NEG | EXTRA | 0.000(0.067) | 0.000(0.206) | 0.56 | 1.000 | $-1.44(3.18)\times10^{-2}$ | -0.079 | -0.45 | 0.991 |
| | TLE- NEG | IGE | 0.000(0.067) | 0.027(0.187) | 1.00 | 1.000 | $2.87(3.16)\times10^{-2}$ | 0.159 | 0.91 | 0.886 |
| | EXTRA | IGE | 0.000(0.206) | 0.027(0.187) | 1.52 | 1.000 | $4.30(3.20)\times10^{-2}$ | 0.238 | 1.35 | 0.644 |
| WM - Parietal | Control | TLE-HS | -0.032(0.214) | 0.000(0.227) | 0.75 | 1.000 | $-7.36(15.50)\times10^{-3}$ | -0.037 | -0.47 | 0.989 |
| | Control | TLE- NEG | -0.032(0.214) | 0.000(0.078) | 2.76 | 0.058 | $4.66(2.55)\times10^{-2}$ | 0.234 | 1.83 | 0.339 |
| | Control | EXTRA | -0.032(0.214) | -0.016(0.157) | 0.43 | 1.000 | $-3.83(26.80)\times10^{-3}$ | -0.019 | -0.14 | 1.000 |
| | Control | IGE | -0.032(0.214) | 0.000(0.220) | 0.02 | 1.000 | $-7.13(26.70)\times10^{-3}$ | -0.036 | -0.27 | 0.999 |
| | TLE-HS | TLE- NEG | 0.000(0.227) | 0.000(0.078) | 2.20 | 0.250 | $5.40(2.66)\times10^{-2}$ | 0.271 | 2.03 | 0.236 |
| | TLE-HS | EXTRA | 0.000(0.227) | -0.016(0.157) | 0.00 | 1.000 | $3.53(28.60)\times10^{-3}$ | 0.018 | 0.12 | 1.000 |
| | TLE-HS | IGE | 0.000(0.227) | 0.000(0.220) | 0.43 | 1.000 | $2.26(283.00)\times10^{-4}$ | 0.001 | 0.01 | 1.000 |
| | TLE- NEG | EXTRA | 0.000(0.078) | -0.016(0.157) | 1.71 | 0.606 | $-5.04(3.51)\times10^{-2}$ | -0.253 | -1.44 | 0.582 |
| | TLE- NEG | IGE | 0.000(0.078) | 0.000(0.220) | 2.05 | 0.321 | $-5.37(3.48)\times10^{-2}$ | -0.270 | -1.54 | 0.514 |
| | EXTRA | IGE | -0.016(0.157) | 0.000(0.220) | 0.34 | 1.000 | $-3.30(35.20)\times10^{-3}$ | -0.017 | -0.09 | 1.000 |
| WM - Temporal | Control | TLE-HS | 0.000(0.444) | 0.000(0.551) | 0.23 | 1.000 | $1.34(40.90)\times10^{-3}$ | 0.003 | 0.03 | 1.000 |
| | Control | TLE- NEG | 0.000(0.444) | 0.000(0.113) | 1.50 | 1.000 | $9.64(6.73)\times10^{-2}$ | 0.184 | 1.43 | 0.586 |
| | Control | EXTRA | 0.000(0.444) | 0.000(0.533) | 1.12 | 1.000 | $9.02(7.07)\times10^{-2}$ | 0.172 | 1.28 | 0.689 |
| | Control | IGE | 0.000(0.444) | 0.000(0.467) | 0.63 | 1.000 | $2.71(7.04)\times10^{-2}$ | 0.052 | 0.39 | 0.995 |
| | TLE-HS | TLE- NEG | 0.000(0.551) | 0.000(0.113) | 1.30 | 1.000 | $9.51(7.01)\times10^{-2}$ | 0.181 | 1.36 | 0.637 |
| | TLE-HS | EXTRA | 0.000(0.551) | 0.000(0.533) | 0.95 | 1.000 | $8.89(7.53)\times10^{-2}$ | 0.169 | 1.18 | 0.747 |
| | TLE-HS | IGE | 0.000(0.551) | 0.000(0.467) | 0.48 | 1.000 | $2.58(7.46)\times10^{-2}$ | 0.049 | 0.35 | 0.997 |
| | TLE- NEG | EXTRA | 0.000(0.113) | 0.000(0.533) | 0.25 | 1.000 | $-6.16(92.40)\times10^{-3}$ | -0.012 | -0.07 | 1.000 |
| | TLE- NEG | IGE | 0.000(0.113) | 0.000(0.467) | 0.62 | 1.000 | $-6.93(9.18)\times10^{-2}$ | -0.132 | -0.75 | 0.938 |
| | EXTRA | IGE | 0.000(0.533) | 0.000(0.467) | 0.36 | 1.000 | $-6.31(9.29)\times10^{-2}$ | -0.120 | -0.68 | 0.957 |
| WM - Occipital | Control | TLE-HS | 0.000(0.667) | 0.000(0.000) | 0.74 | 1.000 | $-4.97(4.63)\times10^{-2}$ | -0.084 | -1.08 | 0.806 |
| | Control | TLE- NEG | 0.000(0.667) | 0.000(0.000) | 0.32 | 1.000 | $-3.09(7.62)\times10^{-2}$ | -0.052 | -0.41 | 0.994 |
| | Control | EXTRA | 0.000(0.667) | 0.000(0.500) | 1.88 | 0.602 | $-16.90(8.01)\times10^{-2}$ | -0.285 | -2.11 | 0.200 |
| | Control | IGE | 0.000(0.667) | 0.000(0.000) | 0.46 | 1.000 | $-4.17(7.97)\times10^{-2}$ | -0.070 | -0.52 | 0.984 |
| | TLE-HS | TLE- NEG | 0.000(0.000) | 0.000(0.000) | 0.12 | 1.000 | $1.88(7.94)\times10^{-2}$ | 0.032 | 0.24 | 0.999 |
| | TLE-HS | EXTRA | 0.000(0.000) | 0.000(0.500) | 1.39 | 1.000 | $-11.90(8.52)\times10^{-2}$ | -0.201 | -1.40 | 0.607 |
| | TLE-HS | IGE | 0.000(0.000) | 0.000(0.000) | 0.03 | 1.000 | $8.06(84.50)\times10^{-3}$ | 0.014 | 0.10 | 1.000 |
| | TLE- NEG | EXTRA | 0.000(0.000) | 0.000(0.500) | 1.21 | 1.000 | $-1.38(1.05)\times10^{-1}$ | -0.233 | -1.32 | 0.659 |
| | TLE- NEG | IGE | 0.000(0.000) | 0.000(0.000) | 0.12 | 1.000 | $-1.08(10.40)\times10^{-2}$ | -0.018 | -0.10 | 1.000 |
| | EXTRA | IGE | 0.000(0.500) | 0.000(0.000) | 1.06 | 1.000 | $1.27(1.05)\times10^{-1}$ | 0.214 | 1.21 | 0.728 |

Table S5.1: PVS Volume Fraction absolute asymmetry in the white matter (WM)

| | Group | | median (IQR) | | Univariate | | General Linear Model | | | | |
|---|---|---|---|---|---|---|---|---|---|---|---|
| | G1 | G2 | G1 | G2 | Dunn | p-Holm | beta(sderr) | d(G) | d(γ) | z | p-Tukey |
| WM | Control | TLE-HS | 0.083(0.121) | 0.075(0.106) | 1.85 | 0.321 | $1.86(7.06)\times10^{-1}$ | -0.048 | 0.2 | 0.26 | 0.999 |
| | Control | TLE- NEG | 0.083(0.121) | 0.041(0.095) | 5.24 | **$1.64\times10^{-6}$** | $7.78(2.01)\times10^{+0}$ | -0.491 | 8.3 | 3.88 | **$9.07\times10^{-4}$** |
| | Control | ETLE | 0.083(0.121) | 0.085(0.099) | 0.99 | 0.971 | $3.96(10.90)\times10^{-1}$ | -0.054 | 0.4 | 0.36 | 0.996 |
| | Control | IGE | 0.083(0.121) | 0.077(0.116) | 0.84 | 0.971 | $2.32(1.31)\times10^{+0}$ | -0.276 | 2.5 | 1.77 | 0.359 |
| | TLE-HS | TLE- NEG | 0.075(0.106) | 0.041(0.095) | 3.93 | **$6.80\times10^{-4}$** | $7.59(2.04)\times10^{+0}$ | -0.443 | 8.1 | 3.72 | 0.001 |
| | TLE-HS | ETLE | 0.075(0.106) | 0.085(0.099) | 1.97 | 0.294 | $2.10(12.10)\times10^{-1}$ | -0.006 | 0.2 | 0.17 | 1.000 |
| | TLE-HS | IGE | 0.075(0.106) | 0.077(0.116) | 0.21 | 0.971 | $2.13(1.40)\times10^{+0}$ | -0.227 | 2.3 | 1.53 | 0.513 |
| | TLE- NEG | ETLE | 0.041(0.095) | 0.085(0.099) | 4.64 | **$3.09\times10^{-5}$** | $-7.38(2.23)\times10^{+0}$ | 0.437 | -7.8 | -3.31 | **0.007** |
| | TLE- NEG | IGE | 0.041(0.095) | 0.077(0.116) | 3.22 | 0.009 | $-5.46(2.34)\times10^{+0}$ | 0.215 | -5.8 | -2.33 | 0.119 |
| | ETLE | IGE | 0.085(0.099) | 0.077(0.116) | 1.38 | 0.673 | $1.92(1.61)\times10^{+0}$ | -0.222 | 2.0 | 1.19 | 0.731 |
| WM - Frontal | Control | TLE-HS | 0.122(0.177) | 0.099(0.146) | 1.52 | 0.623 | $5.11(5.28)\times10^{-1}$ | -0.100 | 0.5 | 0.97 | 0.855 |
| | Control | TLE- NEG | 0.122(0.177) | 0.037(0.105) | 5.82 | **$5.81\times10^{-8}$** | $5.14(1.43)\times10^{+0}$ | -0.458 | 5.1 | 3.60 | **0.003** |
| | Control | ETLE | 0.122(0.177) | 0.139(0.150) | 1.54 | 0.623 | $-2.56(7.17)\times10^{-1}$ | 0.063 | -0.3 | -0.36 | 0.996 |
| | Control | IGE | 0.122(0.177) | 0.128(0.155) | 0.39 | 0.783 | $9.14(8.60)\times10^{-1}$ | -0.170 | 0.9 | 1.06 | 0.807 |
| | TLE-HS | TLE- NEG | 0.099(0.146) | 0.037(0.105) | 4.68 | **$2.30\times10^{-5}$** | $4.63(1.46)\times10^{+0}$ | -0.358 | 4.6 | 3.17 | 0.012 |
| | TLE-HS | ETLE | 0.099(0.146) | 0.139(0.150) | 2.31 | 0.126 | $-7.67(8.23)\times10^{-1}$ | 0.163 | -0.8 | -0.93 | 0.871 |
| | TLE-HS | IGE | 0.099(0.146) | 0.128(0.155) | 1.21 | 0.679 | $4.03(9.41)\times10^{-1}$ | -0.070 | 0.4 | 0.43 | 0.992 |
| | TLE- NEG | ETLE | 0.037(0.105) | 0.139(0.150) | 5.50 | **$3.42\times10^{-7}$** | $-5.39(1.56)\times10^{+0}$ | 0.520 | -5.4 | -3.45 | **0.004** |
| | TLE- NEG | IGE | 0.037(0.105) | 0.128(0.155) | 4.60 | **$2.93\times10^{-5}$** | $-4.22(1.63)\times10^{+0}$ | 0.288 | -4.2 | -2.60 | 0.062 |
| | ETLE | IGE | 0.139(0.150) | 0.128(0.155) | 0.86 | 0.783 | $1.17(1.05)\times10^{+0}$ | -0.232 | 1.2 | 1.12 | 0.777 |
| WM - Parietal | Control | TLE-HS | 0.135(0.188) | 0.124(0.178) | 0.87 | 1.000 | $-4.01(42.30)\times10^{-2}$ | -0.004 | -0.0 | -0.09 | 1.000 |
| | Control | TLE- NEG | 0.135(0.188) | 0.070(0.157) | 4.75 | **$1.87\times10^{-5}$** | $3.80(1.11)\times10^{+0}$ | -0.420 | 4.0 | 3.41 | **0.005** |
| | Control | ETLE | 0.135(0.188) | 0.115(0.221) | 0.43 | 1.000 | $8.64(7.72)\times10^{-1}$ | -0.161 | 0.9 | 1.12 | 0.777 |
| | Control | IGE | 0.135(0.188) | 0.173(0.210) | 1.94 | 0.259 | $-8.47(5.77)\times10^{-1}$ | 0.218 | -0.9 | -1.47 | 0.553 |
| | TLE-HS | TLE- NEG | 0.124(0.178) | 0.070(0.157) | 4.02 | **$4.60\times10^{-4}$** | $3.84(1.13)\times10^{+0}$ | -0.416 | 4.1 | 3.39 | 0.005 |
| | TLE-HS | ETLE | 0.124(0.178) | 0.115(0.221) | 0.08 | 1.000 | $9.04(8.25)\times10^{-1}$ | -0.158 | 1.0 | 1.10 | 0.790 |
| | TLE-HS | IGE | 0.124(0.178) | 0.173(0.210) | 2.34 | 0.115 | $-8.07(6.40)\times10^{-1}$ | 0.222 | -0.9 | -1.26 | 0.690 |
| | TLE- NEG | ETLE | 0.070(0.157) | 0.115(0.221) | 3.19 | **0.010** | $-2.94(1.32)\times10^{+0}$ | 0.258 | -3.1 | -2.22 | 0.153 |
| | TLE- NEG | IGE | 0.070(0.157) | 0.173(0.210) | 5.01 | **$5.52\times10^{-6}$** | $-4.65(1.21)\times10^{+0}$ | 0.638 | -4.9 | -3.83 | **0.001** |
| | ETLE | IGE | 0.115(0.221) | 0.173(0.210) | 1.79 | 0.293 | $-17.10(8.97)\times10^{-1}$ | 0.379 | -1.8 | -1.91 | 0.287 |
| WM - Temporal | Control | TLE-HS | 0.376(0.696) | 0.411(0.695) | 0.69 | 1.000 | $-1.62(1.45)\times10^{-1}$ | 0.085 | -0.2 | -1.12 | 0.778 |
| | Control | TLE- NEG | 0.376(0.696) | 0.066(0.602) | 4.21 | **$2.26\times10^{-4}$** | $11.60(3.63)\times10^{-1}$ | -0.398 | 1.3 | 3.19 | **0.011** |
| | Control | ETLE | 0.376(0.696) | 0.343(0.798) | 0.80 | 1.000 | $1.72(2.65)\times10^{-1}$ | -0.093 | 0.2 | 0.65 | 0.962 |
| | Control | IGE | 0.376(0.696) | 0.386(0.992) | 0.08 | 1.000 | $-9.02(23.70)\times10^{-2}$ | 0.053 | -0.1 | -0.38 | 0.995 |
| | TLE-HS | TLE- NEG | 0.411(0.695) | 0.066(0.602) | 4.41 | **$1.01\times10^{-4}$** | $13.20(3.67)\times10^{-1}$ | -0.483 | 1.5 | 3.59 | **0.003** |
| | TLE-HS | ETLE | 0.411(0.695) | 0.343(0.798) | 1.15 | 1.000 | $3.34(2.80)\times10^{-1}$ | -0.178 | 0.4 | 1.19 | 0.735 |
| | TLE-HS | IGE | 0.411(0.695) | 0.386(0.992) | 0.30 | 1.000 | $7.15(25.20)\times10^{-2}$ | -0.032 | 0.1 | 0.28 | 0.998 |
| | TLE- NEG | ETLE | 0.066(0.602) | 0.343(0.798) | 2.51 | 0.084 | $-9.85(4.35)\times10^{-1}$ | 0.305 | -1.1 | -2.27 | 0.140 |
| | TLE- NEG | IGE | 0.066(0.602) | 0.386(0.992) | 3.18 | **0.012** | $-12.50(4.17)\times10^{-1}$ | 0.451 | -1.5 | -2.99 | **0.020** |
| | ETLE | IGE | 0.343(0.798) | 0.386(0.992) | 0.66 | 1.000 | $-2.63(3.32)\times10^{-1}$ | 0.146 | -0.3 | -0.79 | 0.925 |
| WM - Occipital | Control | TLE-HS | 0.482(0.953) | 0.066(1.000) | 3.99 | **$5.88\times10^{-4}$** | $4.76(1.82)\times10^{-1}$ | -0.227 | 0.5 | 2.62 | 0.059 |
| | Control | TLE- NEG | 0.482(0.953) | 0.000(0.570) | 5.16 | **$2.46\times10^{-6}$** | $17.50(4.64)\times10^{-1}$ | -0.548 | 1.7 | 3.78 | **0.001** |
| | Control | ETLE | 0.482(0.953) | 0.204(1.000) | 1.93 | 0.215 | $4.40(3.18)\times10^{-1}$ | -0.218 | 0.4 | 1.38 | 0.609 |
| | Control | IGE | 0.482(0.953) | 0.143(1.000) | 2.06 | 0.195 | $3.48(3.11)\times10^{-1}$ | -0.176 | 0.3 | 1.12 | 0.776 |
| | TLE-HS | TLE- NEG | 0.066(1.000) | 0.000(0.570) | 2.63 | 0.068 | $12.80(4.78)\times10^{-1}$ | -0.321 | 1.2 | 2.67 | 0.051 |
| | TLE-HS | ETLE | 0.066(1.000) | 0.204(1.000) | 0.36 | 1.000 | $-3.61(34.60)\times10^{-2}$ | 0.009 | -0.0 | -0.10 | 1.000 |
| | TLE-HS | IGE | 0.066(1.000) | 0.143(1.000) | 0.22 | 1.000 | $-1.29(3.39)\times10^{-1}$ | 0.051 | -0.1 | -0.38 | 0.995 |
| | TLE- NEG | ETLE | 0.000(0.570) | 0.204(1.000) | 2.34 | 0.135 | $-13.10(5.50)\times10^{-1}$ | 0.330 | -1.3 | -2.39 | 0.105 |
| | TLE- NEG | IGE | 0.000(0.570) | 0.143(1.000) | 2.22 | 0.160 | $-14.10(5.45)\times10^{-1}$ | 0.372 | -1.4 | -2.58 | 0.065 |
| | ETLE | IGE | 0.204(1.000) | 0.143(1.000) | 0.11 | 1.000 | $-9.26(42.20)\times10^{-2}$ | 0.042 | -0.1 | -0.22 | 0.999 |

Table S5.2: PVS total Volume absolute asymmetry in the white matter (WM)

| | Group | | median (IQR) | | Univariate | | General Linear Model | | | | |
|---|---|---|---|---|---|---|---|---|---|---|---|
| | G1 | G2 | G1 | G2 | Dunn | p-Holm | beta(sderr) | d(G) | d(γ) | z | p-Tukey |
| WM | Control | TLE-HS | 0.082(0.121) | 0.078(0.110) | 0.97 | 0.998 | $-2.98(6.78)\times10^{-1}$ | 0.010 | -0.3 | -0.44 | 0.991 |
| | Control | TLE- NEG | 0.082(0.121) | 0.041(0.096) | 5.02 | **$5.30\times10^{-6}$** | $7.54(1.97)\times10^{+0}$ | -0.475 | 8.0 | 3.83 | **0.001** |
| | Control | ETLE | 0.082(0.121) | 0.088(0.112) | 1.26 | 0.827 | $1.48(10.60)\times10^{-1}$ | -0.020 | 0.2 | 0.14 | 1.000 |
| | Control | IGE | 0.082(0.121) | 0.075(0.118) | 0.89 | 0.998 | $2.36(1.31)\times10^{+0}$ | -0.273 | 2.5 | 1.80 | 0.345 |
| | TLE-HS | TLE- NEG | 0.078(0.110) | 0.041(0.096) | 4.23 | **$1.91\times10^{-4}$** | $7.84(1.99)\times10^{+0}$ | -0.485 | 8.4 | 3.93 | **$7.58\times10^{-4}$** |
| | TLE-HS | ETLE | 0.078(0.110) | 0.088(0.112) | 1.74 | 0.488 | $4.46(11.70)\times10^{-1}$ | -0.030 | 0.5 | 0.38 | 0.995 |
| | TLE-HS | IGE | 0.078(0.110) | 0.075(0.118) | 0.32 | 0.998 | $2.66(1.39)\times10^{+0}$ | -0.283 | 2.8 | 1.92 | 0.281 |
| | TLE- NEG | ETLE | 0.041(0.096) | 0.088(0.112) | 4.69 | **$2.45\times10^{-5}$** | $-7.39(2.18)\times10^{+0}$ | 0.455 | -7.9 | -3.38 | **0.005** |
| | TLE- NEG | IGE | 0.041(0.096) | 0.075(0.118) | 3.01 | **0.018** | $-5.18(2.31)\times10^{+0}$ | 0.202 | -5.5 | -2.24 | 0.145 |
| | ETLE | IGE | 0.088(0.112) | 0.075(0.118) | 1.62 | 0.524 | $2.21(1.60)\times10^{+0}$ | -0.254 | 2.4 | 1.38 | 0.608 |
| WM - Frontal | Control | TLE-HS | 0.113(0.170) | 0.102(0.146) | 1.42 | 0.626 | $4.76(5.26)\times10^{-1}$ | -0.094 | 0.5 | 0.91 | 0.882 |
| | Control | TLE- NEG | 0.113(0.170) | 0.045(0.105) | 5.78 | **$7.56\times10^{-8}$** | $5.15(1.43)\times10^{+0}$ | -0.457 | 5.1 | 3.60 | **0.002** |
| | Control | ETLE | 0.113(0.170) | 0.128(0.178) | 1.67 | 0.472 | $-3.00(7.13)\times10^{-1}$ | 0.073 | -0.3 | -0.42 | 0.992 |
| | Control | IGE | 0.113(0.170) | 0.126(0.159) | 0.55 | 0.799 | $8.11(8.49)\times10^{-1}$ | -0.153 | 0.8 | 0.96 | 0.861 |
| | TLE-HS | TLE- NEG | 0.102(0.146) | 0.045(0.105) | 4.69 | **$2.13\times10^{-5}$** | $4.68(1.46)\times10^{+0}$ | -0.363 | 4.6 | 3.20 | **0.010** |
| | TLE-HS | ETLE | 0.102(0.146) | 0.128(0.178) | 2.38 | 0.103 | $-7.77(8.17)\times10^{-1}$ | 0.168 | -0.8 | -0.95 | 0.863 |
| | TLE-HS | IGE | 0.102(0.146) | 0.126(0.159) | 1.30 | 0.626 | $3.35(9.29)\times10^{-1}$ | -0.058 | 0.3 | 0.36 | 0.996 |
| | TLE- NEG | ETLE | 0.045(0.105) | 0.128(0.178) | 5.57 | **$2.26\times10^{-7}$** | $-5.45(1.56)\times10^{+0}$ | 0.531 | -5.4 | -3.49 | **0.004** |
| | TLE- NEG | IGE | 0.045(0.105) | 0.126(0.159) | 4.69 | **$2.13\times10^{-5}$** | $-4.34(1.62)\times10^{+0}$ | 0.305 | -4.3 | -2.67 | 0.050 |
| | ETLE | IGE | 0.128(0.178) | 0.126(0.159) | 0.84 | 0.799 | $1.11(1.03)\times10^{+0}$ | -0.226 | 1.1 | 1.07 | 0.801 |
| WM - Parietal | Control | TLE-HS | 0.137(0.183) | 0.114(0.180) | 0.96 | 1.000 | $-6.86(43.00)\times10^{-2}$ | 0.002 | -0.1 | -0.16 | 1.000 |
| | Control | TLE- NEG | 0.137(0.183) | 0.073(0.145) | 4.59 | **$3.95\times10^{-5}$** | $3.96(1.15)\times10^{+0}$ | -0.422 | 4.1 | 3.44 | **0.005** |
| | Control | ETLE | 0.137(0.183) | 0.120(0.221) | 0.32 | 1.000 | $7.61(7.80)\times10^{-1}$ | -0.140 | 0.8 | 0.98 | 0.852 |
| | Control | IGE | 0.137(0.183) | 0.161(0.211) | 1.86 | 0.311 | $-8.80(5.88)\times10^{-1}$ | 0.222 | -0.9 | -1.50 | 0.535 |
| | TLE-HS | TLE- NEG | 0.114(0.180) | 0.073(0.145) | 3.82 | **0.001** | $4.03(1.17)\times10^{+0}$ | -0.424 | 4.2 | 3.44 | **0.005** |
| | TLE-HS | ETLE | 0.114(0.180) | 0.120(0.221) | 0.23 | 1.000 | $8.30(8.33)\times10^{-1}$ | -0.143 | 0.9 | 1.00 | 0.842 |
| | TLE-HS | IGE | 0.114(0.180) | 0.161(0.211) | 2.31 | 0.124 | $-8.11(6.51)\times10^{-1}$ | 0.219 | -0.8 | -1.25 | 0.700 |
| | TLE- NEG | ETLE | 0.073(0.145) | 0.120(0.221) | 3.16 | **0.011** | $-3.20(1.35)\times10^{+0}$ | 0.282 | -3.3 | -2.36 | 0.111 |
| | TLE- NEG | IGE | 0.073(0.145) | 0.161(0.211) | 4.83 | **$1.35\times10^{-5}$** | $-4.84(1.25)\times10^{+0}$ | 0.644 | -5.1 | -3.87 | **$7.94\times10^{-4}$** |
| | ETLE | IGE | 0.120(0.221) | 0.161(0.211) | 1.65 | 0.394 | $-16.40(9.08)\times10^{-1}$ | 0.362 | -1.7 | -1.81 | 0.341 |
| WM - Temporal | Control | TLE-HS | 0.379(0.698) | 0.412(0.682) | 0.76 | 1.000 | $-1.65(1.45)\times10^{-1}$ | 0.087 | -0.2 | -1.14 | 0.765 |
| | Control | TLE- NEG | 0.379(0.698) | 0.083(0.618) | 4.15 | **$3.01\times10^{-4}$** | $11.80(3.66)\times10^{-1}$ | -0.402 | 1.4 | 3.22 | **0.010** |
| | Control | ETLE | 0.379(0.698) | 0.333(0.833) | 0.78 | 1.000 | $1.71(2.67)\times10^{-1}$ | -0.091 | 0.2 | 0.64 | 0.964 |
| | Control | IGE | 0.379(0.698) | 0.381(1.000) | 0.16 | 1.000 | $-9.59(23.80)\times10^{-2}$ | 0.057 | -0.1 | -0.40 | 0.994 |
| | TLE-HS | TLE- NEG | 0.412(0.682) | 0.083(0.618) | 4.39 | **$1.13\times10^{-4}$** | $13.40(3.71)\times10^{-1}$ | -0.489 | 1.6 | 3.63 | **0.002** |
| | TLE-HS | ETLE | 0.412(0.682) | 0.333(0.833) | 1.17 | 1.000 | $3.36(2.81)\times10^{-1}$ | -0.179 | 0.4 | 1.19 | 0.733 |
| | TLE-HS | IGE | 0.412(0.682) | 0.381(1.000) | 0.27 | 1.000 | $6.95(25.30)\times10^{-2}$ | -0.031 | 0.1 | 0.27 | 0.999 |
| | TLE- NEG | ETLE | 0.083(0.618) | 0.333(0.833) | 2.47 | 0.094 | $-10.10(4.38)\times10^{-1}$ | 0.310 | -1.2 | -2.30 | 0.129 |
| | TLE- NEG | IGE | 0.083(0.618) | 0.381(1.000) | 3.19 | **0.012** | $-12.80(4.21)\times10^{-1}$ | 0.458 | -1.5 | -3.03 | **0.018** |
| | ETLE | IGE | 0.333(0.833) | 0.381(1.000) | 0.71 | 1.000 | $-2.67(3.33)\times10^{-1}$ | 0.148 | -0.3 | -0.80 | 0.922 |
| WM - Occipital | Control | TLE-HS | 0.500(0.949) | 0.048(1.000) | 3.84 | **0.001** | $4.73(1.82)\times10^{-1}$ | -0.226 | 0.5 | 2.60 | 0.062 |
| | Control | TLE- NEG | 0.500(0.949) | 0.000(0.536) | 5.07 | **$4.07\times10^{-6}$** | $17.90(4.68)\times10^{-1}$ | -0.554 | 1.7 | 3.82 | **0.001** |
| | Control | ETLE | 0.500(0.949) | 0.188(1.000) | 1.78 | 0.300 | $4.45(3.19)\times10^{-1}$ | -0.220 | 0.4 | 1.39 | 0.602 |
| | Control | IGE | 0.500(0.949) | 0.100(1.000) | 1.94 | 0.261 | $3.63(3.13)\times10^{-1}$ | -0.183 | 0.4 | 1.16 | 0.752 |
| | TLE-HS | TLE- NEG | 0.048(1.000) | 0.000(0.536) | 2.63 | 0.069 | $13.10(4.83)\times10^{-1}$ | -0.328 | 1.3 | 2.72 | **0.044** |
| | TLE-HS | ETLE | 0.048(1.000) | 0.188(1.000) | 0.42 | 1.000 | $-2.89(34.70)\times10^{-2}$ | 0.006 | -0.0 | -0.08 | 1.000 |
| | TLE-HS | IGE | 0.048(1.000) | 0.100(1.000) | 0.26 | 1.000 | $-1.10(3.41)\times10^{-1}$ | 0.043 | -0.1 | -0.32 | 0.997 |
| | TLE- NEG | ETLE | 0.000(0.536) | 0.188(1.000) | 2.38 | 0.120 | $-13.40(5.54)\times10^{-1}$ | 0.334 | -1.3 | -2.42 | 0.096 |
| | TLE- NEG | IGE | 0.000(0.536) | 0.100(1.000) | 2.24 | 0.150 | $-14.20(5.50)\times10^{-1}$ | 0.371 | -1.4 | -2.59 | 0.063 |
| | ETLE | IGE | 0.188(1.000) | 0.100(1.000) | 0.13 | 1.000 | $-8.15(42.40)\times10^{-2}$ | 0.037 | -0.1 | -0.19 | 1.000 |

Table S5.3: PVS number absolute asymmetry in the white matter (WM)

| | Group | | median (IQR) | | Univariate | | General Linear Model | | | | |
|---|---|---|---|---|---|---|---|---|---|---|---|
| | G1 | G2 | G1 | G2 | Dunn | p-Holm | beta(sderr) | d(G) | d(γ) | z | p-Tukey |
| WM | Control | TLE-HS | 0.067(0.095) | 0.071(0.094) | 0.36 | 1.000 | -2.31(7.90)x10$^{-1}$ | 0.004 | -0.3 | -0.29 | 0.998 |
| | Control | TLE- NEG | 0.067(0.095) | 0.018(0.067) | 5.99 | **2.11x10$^{-8}$** | 11.20(2.52)x10$^{+0}$ | -0.559 | 12.2 | 4.44 | **6.12x10$^{-5}$** |
| | Control | ETLE | 0.067(0.095) | 0.073(0.052) | 0.83 | 1.000 | 9.42(13.60)x10$^{-1}$ | -0.107 | 1.0 | 0.69 | 0.951 |
| | Control | IGE | 0.067(0.095) | 0.057(0.058) | 1.45 | 0.590 | 3.68(1.64)x10$^{+0}$ | -0.337 | 4.0 | 2.24 | 0.145 |
| | TLE-HS | TLE- NEG | 0.071(0.094) | 0.018(0.067) | 5.91 | **2.99x10$^{-8}$** | 11.40(2.55)x10$^{+0}$ | -0.562 | 12.4 | 4.49 | **8.92x10$^{-5}$** |
| | TLE-HS | ETLE | 0.071(0.094) | 0.073(0.052) | 0.60 | 1.000 | 1.17(1.47)x10$^{+0}$ | -0.110 | 1.3 | 0.80 | 0.920 |
| | TLE-HS | IGE | 0.071(0.094) | 0.057(0.058) | 1.58 | 0.566 | 3.91(1.72)x10$^{+0}$ | -0.340 | 4.3 | 2.27 | 0.135 |
| | TLE- NEG | ETLE | 0.018(0.067) | 0.073(0.052) | 5.08 | **2.99x10$^{-6}$** | -10.30(2.81)x10$^{+0}$ | 0.452 | -11.2 | -3.66 | **0.002** |
| | TLE- NEG | IGE | 0.018(0.067) | 0.057(0.058) | 3.30 | **0.007** | -7.53(2.95)x10$^{+0}$ | 0.222 | -8.2 | -2.55 | 0.068 |
| | ETLE | IGE | 0.073(0.052) | 0.057(0.058) | 1.72 | 0.512 | 2.74(2.03)x10$^{+0}$ | -0.230 | 3.0 | 1.35 | 0.630 |
| WM - Frontal | Control | TLE-HS | 0.100(0.140) | 0.075(0.122) | 2.80 | 0.031 | 14.10(6.53)x10$^{-1}$ | -0.198 | 1.4 | 2.15 | 0.175 |
| | Control | TLE- NEG | 0.100(0.140) | 0.030(0.093) | 6.32 | **2.67x10$^{-9}$** | 8.10(1.90)x10$^{+0}$ | -0.570 | 8.1 | 4.25 | **1.70x10$^{-4}$** |
| | Control | ETLE | 0.100(0.140) | 0.097(0.152) | 0.12 | 1.000 | 7.64(9.66)x10$^{-1}$ | -0.126 | 0.8 | 0.79 | 0.924 |
| | Control | IGE | 0.100(0.140) | 0.093(0.107) | 0.49 | 1.000 | 2.07(1.12)x10$^{+0}$ | -0.297 | 2.1 | 1.85 | 0.317 |
| | TLE-HS | TLE- NEG | 0.075(0.122) | 0.030(0.093) | 4.42 | **8.06x10$^{-5}$** | 6.69(1.95)x10$^{+0}$ | -0.371 | 6.7 | 3.44 | **0.005** |
| | TLE-HS | ETLE | 0.075(0.122) | 0.097(0.152) | 1.43 | 0.762 | -6.43(11.00)x10$^{-1}$ | 0.072 | -0.6 | -0.59 | 0.974 |
| | TLE-HS | IGE | 0.075(0.122) | 0.093(0.107) | 1.07 | 1.000 | 6.59(12.30)x10$^{-1}$ | -0.099 | 0.7 | 0.54 | 0.981 |
| | TLE- NEG | ETLE | 0.030(0.093) | 0.097(0.152) | 4.59 | **4.03x10$^{-5}$** | -7.33(2.10)x10$^{+0}$ | 0.444 | -7.3 | -3.50 | **0.004** |
| | TLE- NEG | IGE | 0.030(0.093) | 0.093(0.107) | 4.28 | **1.29x10$^{-4}$** | -6.03(2.17)x10$^{+0}$ | 0.272 | -6.0 | -2.78 | **0.037** |
| | ETLE | IGE | 0.097(0.152) | 0.093(0.107) | 0.28 | 1.000 | 1.30(1.41)x10$^{+0}$ | -0.171 | 1.3 | 0.93 | 0.872 |
| WM - Parietal | Control | TLE-HS | 0.111(0.145) | 0.111(0.144) | 0.51 | 1.000 | -2.75(5.35)x10$^{-1}$ | 0.018 | -0.3 | -0.52 | 0.984 |
| | Control | TLE- NEG | 0.111(0.145) | 0.053(0.123) | 5.34 | **9.04x10$^{-7}$** | 5.78(1.57)x10$^{+0}$ | -0.454 | 5.9 | 3.67 | **0.002** |
| | Control | ETLE | 0.111(0.145) | 0.083(0.102) | 1.72 | 0.510 | 1.64(1.03)x10$^{+0}$ | -0.237 | 1.7 | 1.59 | 0.472 |
| | Control | IGE | 0.111(0.145) | 0.111(0.189) | 0.24 | 1.000 | -1.50(8.14)x10$^{-1}$ | 0.020 | -0.2 | -0.18 | 1.000 |
| | TLE-HS | TLE- NEG | 0.111(0.144) | 0.053(0.123) | 4.80 | **1.40x10$^{-5}$** | 6.06(1.59)x10$^{+0}$ | -0.472 | 6.1 | 3.80 | **0.001** |
| | TLE-HS | ETLE | 0.111(0.144) | 0.083(0.102) | 1.37 | 0.860 | 1.92(1.10)x10$^{+0}$ | -0.255 | 1.9 | 1.75 | 0.374 |
| | TLE-HS | IGE | 0.111(0.144) | 0.111(0.189) | 0.05 | 1.000 | 1.26(8.87)x10$^{-1}$ | 0.003 | 0.1 | 0.14 | 1.000 |
| | TLE- NEG | ETLE | 0.053(0.123) | 0.083(0.102) | 2.64 | 0.059 | -4.14(1.84)x10$^{+0}$ | 0.217 | -4.2 | -2.25 | 0.144 |
| | TLE- NEG | IGE | 0.053(0.123) | 0.111(0.189) | 3.76 | **0.001** | -5.93(1.73)x10$^{+0}$ | 0.475 | -6.0 | -3.44 | **0.004** |
| | ETLE | IGE | 0.083(0.102) | 0.111(0.189) | 1.11 | 1.000 | -1.79(1.24)x10$^{+0}$ | 0.258 | -1.8 | -1.44 | 0.568 |
| WM - Temporal | Control | TLE-HS | 0.227(0.600) | 0.333(0.523) | 1.50 | 0.800 | -2.77(2.09)x10$^{-1}$ | 0.103 | -0.3 | -1.33 | 0.653 |
| | Control | TLE- NEG | 0.227(0.600) | 0.077(0.487) | 2.55 | 0.098 | 9.05(4.72)x10$^{-1}$ | -0.231 | 0.9 | 1.92 | 0.286 |
| | Control | ETLE | 0.227(0.600) | 0.250(0.600) | 0.22 | 1.000 | -1.14(3.61)x10$^{-1}$ | 0.043 | -0.1 | -0.32 | 0.998 |
| | Control | IGE | 0.227(0.600) | 0.333(1.000) | 0.13 | 1.000 | -2.55(3.39)x10$^{-1}$ | 0.102 | -0.2 | -0.75 | 0.938 |
| | TLE-HS | TLE- NEG | 0.333(0.523) | 0.077(0.487) | 3.29 | **0.010** | 11.80(4.78)x10$^{-1}$ | -0.334 | 1.2 | 2.47 | 0.087 |
| | TLE-HS | ETLE | 0.333(0.523) | 0.250(0.600) | 0.62 | 1.000 | 1.63(3.82)x10$^{-1}$ | -0.060 | 0.2 | 0.43 | 0.992 |
| | TLE-HS | IGE | 0.333(0.523) | 0.333(1.000) | 0.70 | 1.000 | 2.21(35.90)x10$^{-2}$ | -0.001 | 0.0 | 0.06 | 1.000 |
| | TLE- NEG | ETLE | 0.077(0.487) | 0.250(0.600) | 2.06 | 0.319 | -10.20(5.69)x10$^{-1}$ | 0.273 | -1.0 | -1.79 | 0.356 |
| | TLE- NEG | IGE | 0.077(0.487) | 0.333(1.000) | 1.98 | 0.331 | -11.60(5.54)x10$^{-1}$ | 0.333 | -1.1 | -2.09 | 0.205 |
| | ETLE | IGE | 0.250(0.600) | 0.333(1.000) | 0.06 | 1.000 | -1.41(4.57)x10$^{-1}$ | 0.059 | -0.1 | -0.31 | 0.998 |
| WM - Occipital | Control | TLE-HS | 0.333(1.000) | 0.000(1.000) | 3.13 | 0.016 | 3.55(2.23)x10$^{-1}$ | -0.135 | 0.3 | 1.60 | 0.471 |
| | Control | TLE- NEG | 0.333(1.000) | 0.000(0.350) | 4.36 | **1.31x10$^{-4}$** | 17.00(5.52)x10$^{-1}$ | -0.426 | 1.5 | 3.07 | **0.016** |
| | Control | ETLE | 0.333(1.000) | 0.000(1.000) | 1.44 | 0.628 | 3.08(3.79)x10$^{-1}$ | -0.122 | 0.3 | 0.81 | 0.918 |
| | Control | IGE | 0.333(1.000) | 0.000(1.000) | 1.53 | 0.628 | 2.59(3.80)x10$^{-1}$ | -0.103 | 0.2 | 0.68 | 0.955 |
| | TLE-HS | TLE- NEG | 0.000(1.000) | 0.000(0.350) | 2.36 | 0.146 | 13.40(5.68)x10$^{-1}$ | -0.291 | 1.2 | 2.36 | 0.112 |
| | TLE-HS | ETLE | 0.000(1.000) | 0.000(1.000) | 0.36 | 1.000 | -4.68(41.20)x10$^{-2}$ | 0.013 | -0.0 | -0.11 | 1.000 |
| | TLE-HS | IGE | 0.000(1.000) | 0.000(1.000) | 0.26 | 1.000 | -9.58(41.10)x10$^{-2}$ | 0.032 | -0.1 | -0.23 | 0.999 |
| | TLE- NEG | ETLE | 0.000(0.350) | 0.000(1.000) | 2.12 | 0.236 | -13.90(6.52)x10$^{-1}$ | 0.304 | -1.2 | -2.13 | 0.187 |
| | TLE- NEG | IGE | 0.000(0.350) | 0.000(1.000) | 2.04 | 0.251 | -14.40(6.52)x10$^{-1}$ | 0.324 | -1.3 | -2.21 | 0.159 |
| | ETLE | IGE | 0.000(1.000) | 0.000(1.000) | 0.08 | 1.000 | -4.90(50.60)x10$^{-2}$ | 0.019 | -0.0 | -0.10 | 1.000 |

Table S6: Association of duration of illness with PVS Volume Fraction in each region and sub-region, corrected for age and sex. (WM: White Matter, BG: basal ganglia).

| | beta (std error) | t | p |
|---|---|---|---|
| WM | -4.49e-06(6.93e-06) | -0.65 | 0.518 |
| WM Frontal | -8.83e-06(1.04e-05) | -0.85 | 0.398 |
| WM Parietal | -5.48e-06(1.24e-05) | -0.44 | 0.658 |
| WM Temporal | -9.15e-07(3.63e-06) | -0.25 | 0.801 |
| WM Occipital | 2.61e-06(1.71e-06) | 1.53 | 0.127 |
| Basal Ganglia | -1.71e-06(1.18e-05) | -0.15 | 0.885 |
| BG exc. Thalami | -3.77e-06(1.52e-05) | -0.25 | 0.803 |
| Thalami | 2.96e-06(3.08e-06) | 0.96 | 0.337 |

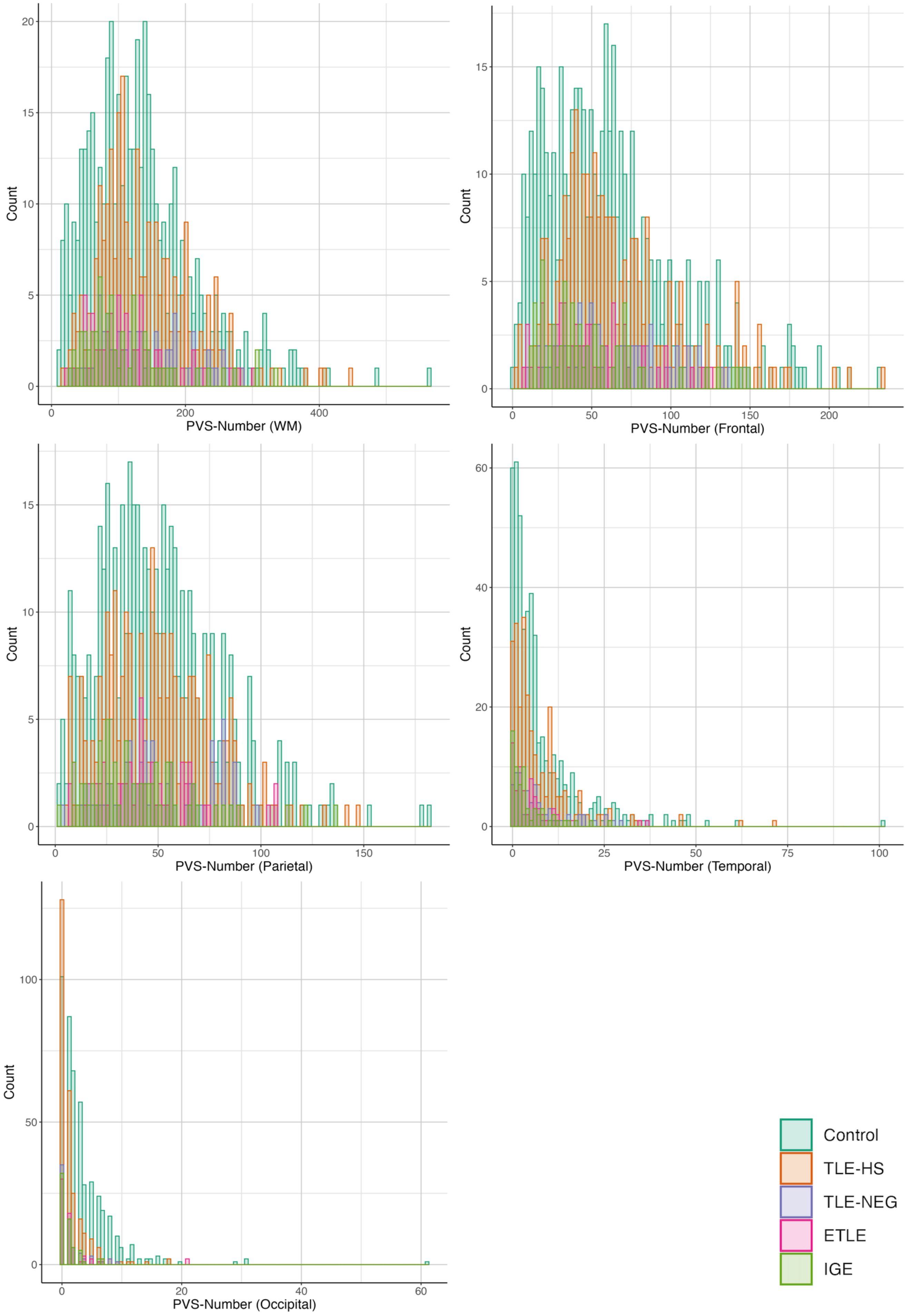

Figure S1: Histograms of number of PVS in each White Matter (WM) region

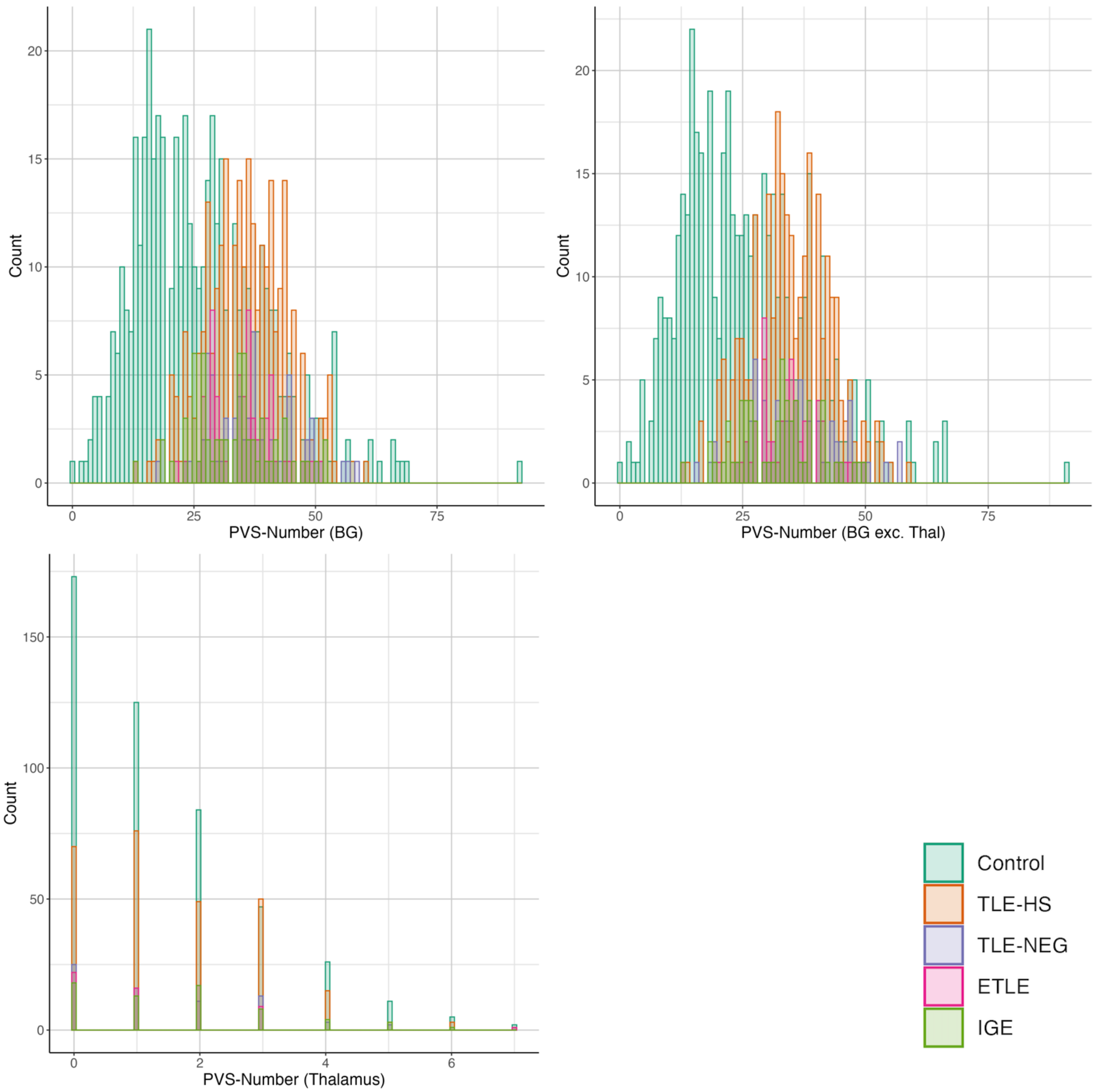

Figure S2: Histograms of number of PVS in each Basal Ganglia (BG) region

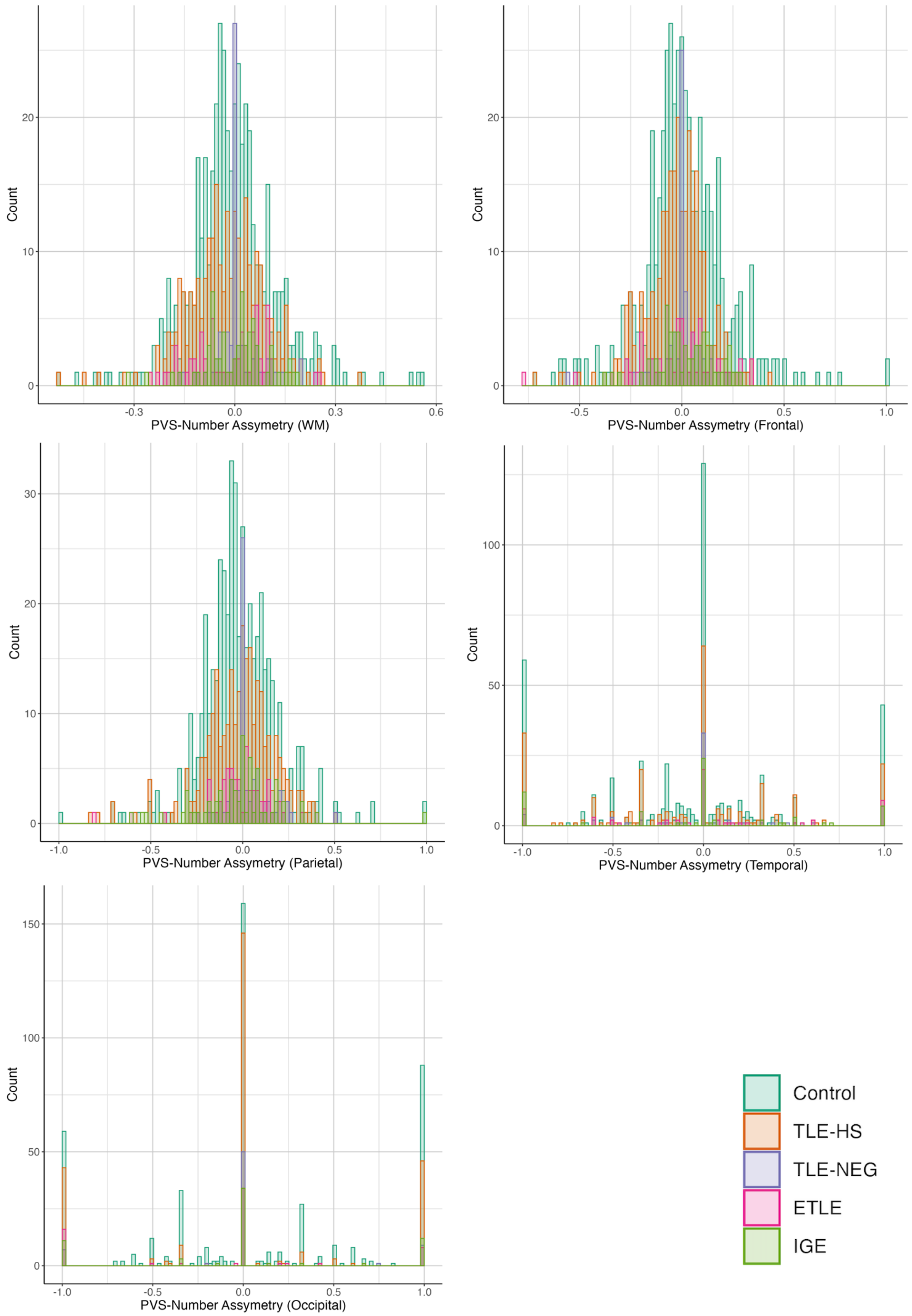

Figure S3: Histograms PVS-Number asymmetry in each White Matter (WM) region

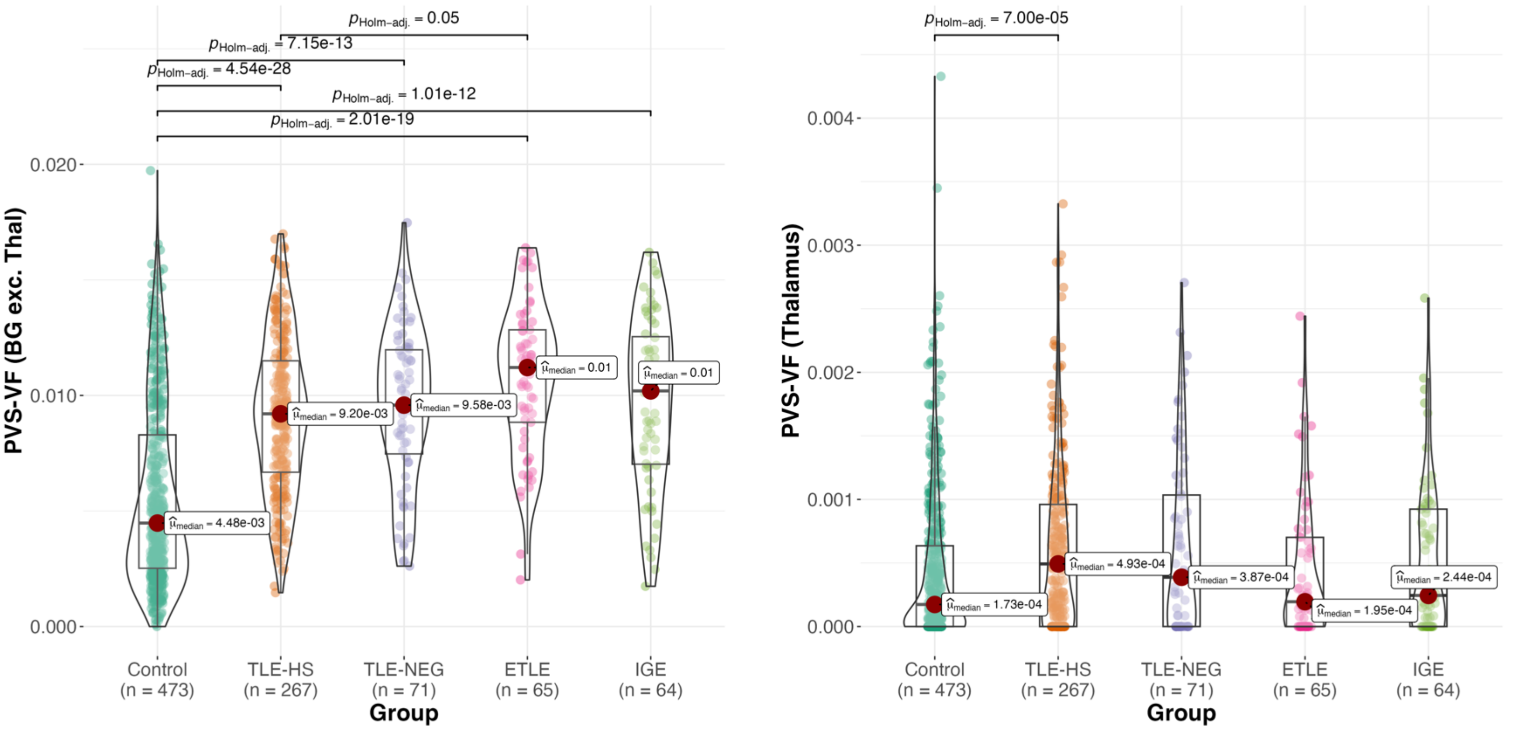

Figure S4: PVS volume fraction in Basal Ganglia excluding Thalami (BG exc. Thalami, left) and Thalami (Thal, right) by group
